# ChatCLIDS: Simulating Persuasive AI Dialogues to Promote Closed-Loop Insulin Adoption in Type 1 Diabetes Care

**DOI:** 10.1101/2025.09.02.25334973

**Authors:** Zonghai Yao, Talha Chafekar, Junda Wang, Shuo Han, Feiyun Ouyang, Junhui Qian, Lingxi Li, Hong Yu

## Abstract

Real-world adoption of closed-loop insulin delivery systems (CLIDS) in type 1 diabetes remains low, driven not by technical failure, but by diverse behavioral, psychosocial, and social barriers. We introduce ChatCLIDS, the first benchmark to rigorously evaluate LLM–driven persuasive dialogue for health behavior change. Our framework features a library of expert-validated virtual patients, each with clinically grounded, heterogeneous profiles and realistic adoption barriers, and simulates multi-turn interactions with nurse agents equipped with a diverse set of evidence-based persuasive strategies. ChatCLIDS uniquely supports longitudinal counseling and adversarial social influence scenarios, enabling robust, multi-dimensional evaluation. Our findings reveal that while larger and more reflective LLMs adapt strategies over time, all models struggle to overcome resistance, especially under realistic social pressure. These results highlight critical limitations of current LLMs for behavior change, and offer a high-fidelity, scalable testbed for advancing trustworthy persuasive AI in healthcare and beyond. ^1^

## Introduction

Type 1 diabetes (T1D) is a lifelong condition that imposes a relentless physical and psychological burden on millions worldwide (CDC 2024). Recent advances such as hybrid closed-loop insulin delivery systems (CLIDS) have transformed the clinical possibilities of diabetes care, offering automated, real-time glucose monitoring and insulin dosing (Manero 2023; Borel et al. 2024). Yet, despite clear medical benefits, real-world adoption of CLIDS remains strikingly low, fewer than 25% of eligible patients initiate use, and up to 30% discontinue within six months (Noor et al. 2022; Messer et al. 2020; Wong et al. 2017).

The reasons for this gap are multifaceted and extend far beyond technical performance. Patients and families face a daunting array of CLIDS devices with different features, algorithms, and user requirements, making device selection and transition confusing and overwhelming (Manero 2023; Saunders, Messer, and Forlenza 2019; Cobry et al. 2020). Technical challenges such as calibration demands and alarm fatigue are common, but psychosocial and behavioral barriers, including reluctance to cede self-management, anxiety about trusting automated systems, emotional discomfort, financial concerns, prior negative experiences, and low confidence in adopting new technology, are often even more decisive (Manero 2023) (Figure 1). Overcoming these obstacles requires not just initial instruction, but ongoing, individualized education, expectation-setting, and emotional support, resources that are difficult to provide at scale in current healthcare systems (Manero 2023; Tanenbaum and Commissariat 2022). Importantly, low adoption of advanced diabetes technologies risks further widening health disparities, as under-resourced and marginalized patients often face the steepest barriers to sustained engagement and benefit.

**Figure 1:**
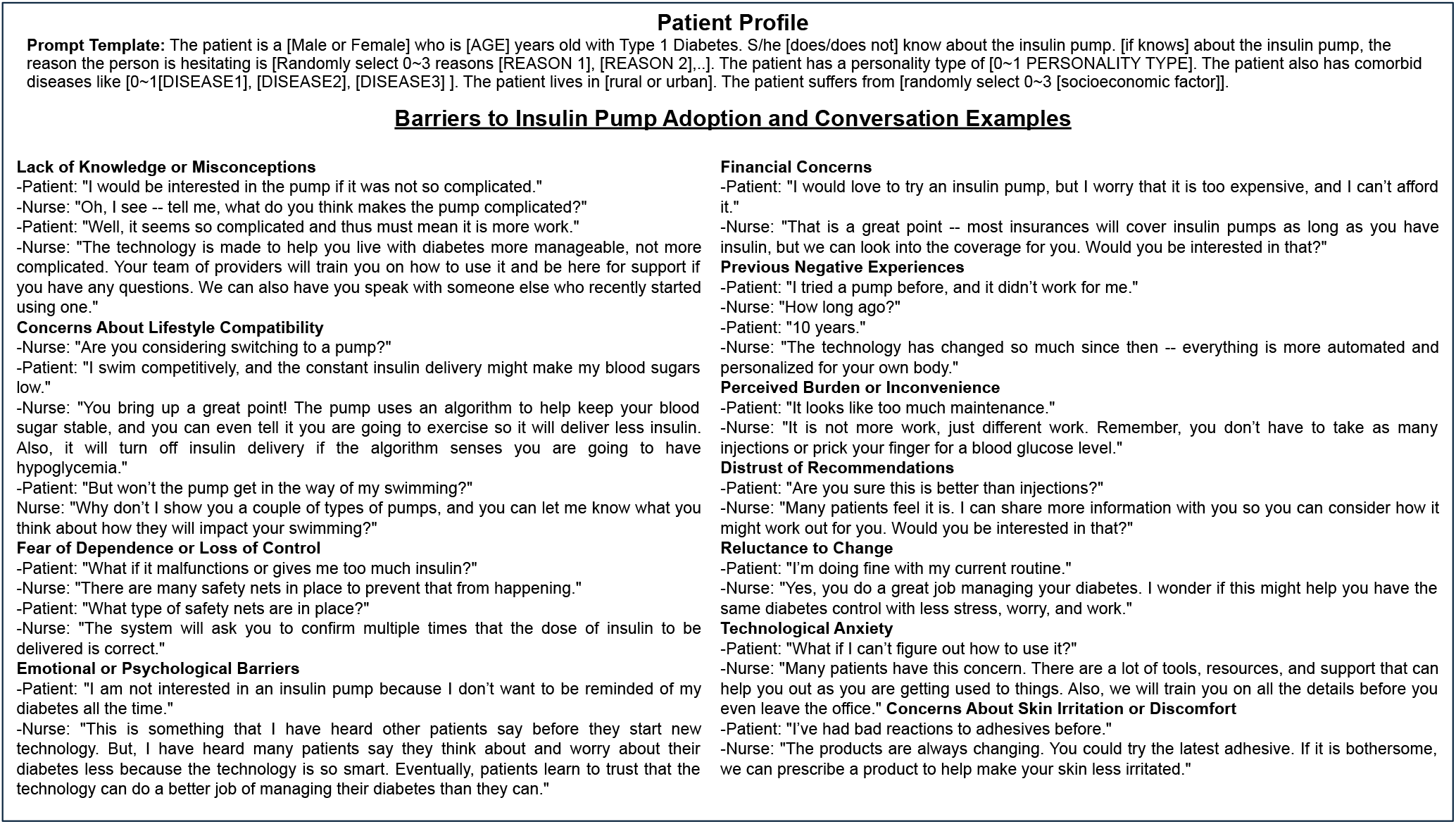
Structure of the Patient Agent in ChatCLIDS. Each agent is initialized with a clinically validated profile and a scenario-driven set of adoption barriers. The resulting diversity in persuasion barriers and conversational responses enables personalized, realistic, and challenging evaluation of persuasive dialogue systems.

To enable rigorous and scalable investigation of real-world behavioral barriers, we introduce a diverse library of virtual patient agents. Each agent is systematically initialized with a clinically validated profile and a set of realistic, scenario-driven adoption barriers, as visualized in Figure 1. This design captures the heterogeneity of attitudes, misconceptions, and behavioral resistance observed among people living with T1D, enabling high-fidelity, customizable, and clinically meaningful evaluation of AI-driven interventions.

To address the complex requirements of this domain, we propose ChatCLIDS, the first benchmark expressly developed to assess LLM-driven persuasive dialogue for behavior change in healthcare. ChatCLIDS centers on structured, multi-turn, and outcome-oriented conversations, with a clinical target: increasing CLIDS adoption among T1D patients. The benchmark features two interacting LLM agents: a Patient Agent, initialized with rigorously curated and expert-validated clinical and psychosocial profiles, and a Nurse Agent, equipped with an extensive repertoire of evidence-based persuasive strategies, including empathy, logical reasoning, expert endorsement, and motivational coaching.

ChatCLIDS supports clinically realistic scenarios of varying difficulty (Easy, Medium, Hard), encompassing both single-visit and multi-visit settings to capture the longitu-dinal dynamics of dialogue. The framework also includes a Social Resistance Agent to model peer pressure and misinformation, mirroring real-world social influences. Evaluation is multi-dimensional and robust: model outputs are assessed on responsiveness, empathy, appropriateness of strategy, clinical relevance, and behavioral realism, using both expert annotation and advanced LLM-based judges (see Appendix LLM-as-Judge Evaluation). The result is a scalable, transparent, and clinically grounded testbed for AI-facilitated patient persuasion, with careful design choices to ensure realism and reliability. In our experiments, we find that:

- **Scaling and Performance:** In single-visit settings, model performance generally scales with model size, but the best “non-reasoning” and “reasoning” models achieve similar results.
- **Challenge of Hard Cases:** All models struggle to persuade “medium” and “hard” patients in a single visit, with some interventions leading to adverse outcomes.
- **Value of Chain of Strategy:** The Chain of Strategy (CoS) protocol markedly boosts most LLMs’ effectiveness on this challenging persuasion task.
- **Role of Reflection and Adaptation:** In multi-visit settings, models endowed with explicit reflection mechanisms (e.g., DeepSeek-R1 (Guo et al. 2025), GPT-o4-mini (Jaech et al. 2024)) exhibit substantial gains over “no thinking” models. These reflective agents learn to select strategies more suited to individual patient barriers and can discard approaches that previously failed.
- **Social Complexity and Failure Modes:** When exposed to realistic, adversarial social influences (the “Social Resistance” scenario), all agents, even the most advanced, experience dramatic performance degradation. Prolonged, multi-visit interventions rarely achieve persuasion within practical timeframes, and the presence of persistent social resistance can even entrench negative attitudes. These findings highlight the gap between simulated persuasion success and the complexity of real-world social environments, indicating that present-day LLMs lack the robustness, resilience, and situational awareness necessary to drive sustained health behavior change at scale.

## Related Works

### LLM Applications for Persuasion

LLMs have recently demonstrated human-level persuasive capabilities across a range of applications, including prosocial messaging and attitude change (Manca et al. 2020; Xiu et al. 2024; Zeng et al. 2024; Gordon 1993). Prior research on persuasion has primarily relied on human studies evaluating message effectiveness (Manca et al. 2020; Xiu et al. 2024), while emerging frameworks, such as PersuasionArena and Convincer-Skeptic, explore agent-based and automated approaches (Zeng et al. 2024; Gordon 1993). However, most of these works focus on single-turn, open-domain, or debate-oriented scenarios (Bozdag et al. 2025), with limited emphasis on sustained, goal-directed behavioral change in real-world health settings. In the clinical context, patient engagement and persuasion are well-recognized as critical strategies for improving health outcomes (Marzban et al. 2022; Geurts et al. 2022). Still, there is a lack of systematic and reproducible benchmarks for studying persuasive dialogue that targets complex medical decisions. ChatCLIDS addresses this gap by modeling cl9inically grounded, multi-turn persuasion between virtual nurse and patient agents, centered on the real-world challenge of CLIDS adoption in T1D care.

### Multi-Agent Simulation Frameworks in Healthcare

Multi-agent simulation has emerged as a key approach for modeling complex, dynamic healthcare interactions involving multiple stakeholders (Yao and Yu 2025; Tariq 2024; Elkamouchi, Daaif, and Elguemmat 2024; Daengdej et al. 2024). Recent advances leverage LLM-based agents to simulate clinical workflows, diagnostic reasoning, and patient-provider communication (Wang et al. 2023; Cai et al. 2023; Zhang et al. 2023). Frameworks such as Agent-Clinic (Schmidgall et al. 2024) and AMIE (Tu et al. 2024) utilize interactive language agents for benchmarking clinical decision-making, scenario generation, and clinician training, supporting adaptive, multimodal interactions. In the context of T1D, however, most prior systems have not addressed the unique behavioral, psychological, and social challenges surrounding health technology adoption. Our work builds on this line of research by introducing a clinically realistic multi-agent framework specifically tailored for simulating T1D patient persuasion and resistance in the context of CLIDS adoption.

### Virtual Patients and Realistic Agent Modeling

Virtual patients have a long history in clinical education, traditionally implemented with rule-based systems to simulate demographic and medical features (Huang, Reynolds, and Candler 2007; Gordon et al. 2001; Good 2003; Garrett, MacPhee, and Jackson 2010; Campillos-Llanos et al. 2021). With the advent of LLMs, recent research has demonstrated the potential for highly realistic, behaviorally nuanced agent simulation, including the accurate reproduction of individual social attitudes, psychological barriers, and conversational dynamics (Lee et al. 2024; Wang et al. 2024; Park et al. 2023). For example, Park et al. (2024) demonstrated that LLM-driven agents can closely replicate the survey responses, personality traits, and longitudinal behaviors of real individuals. In medicine, advanced agent-based platforms enable simulation of both provider and patient roles, facilitating scalable, ethical, and controlled study of decision-making and patient engagement (Schmidgall et al. 2024; Tu et al. 2024; Yu et al. 2024). ChatCLIDS leverages these advancements to develop expert-validated, highly customizable virtual T1D patients, systematically capturing the full spectrum of knowledge, attitudes, comorbidities, and social factors relevant to technology adoption.

## Methodology

ChatCLIDS is a clinically grounded, agent-based benchmark for evaluating the persuasive capacity of LLMs in health behavior change. As illustrated in Figure 1 and Figure 2, the benchmark simulates multi-turn, multi-session conversations between a Nurse Agent and a Patient Agent, under three progressively challenging real-world scenarios: Single-Visit, Multi-Visit, and Social Resistance.

**Figure 2:**
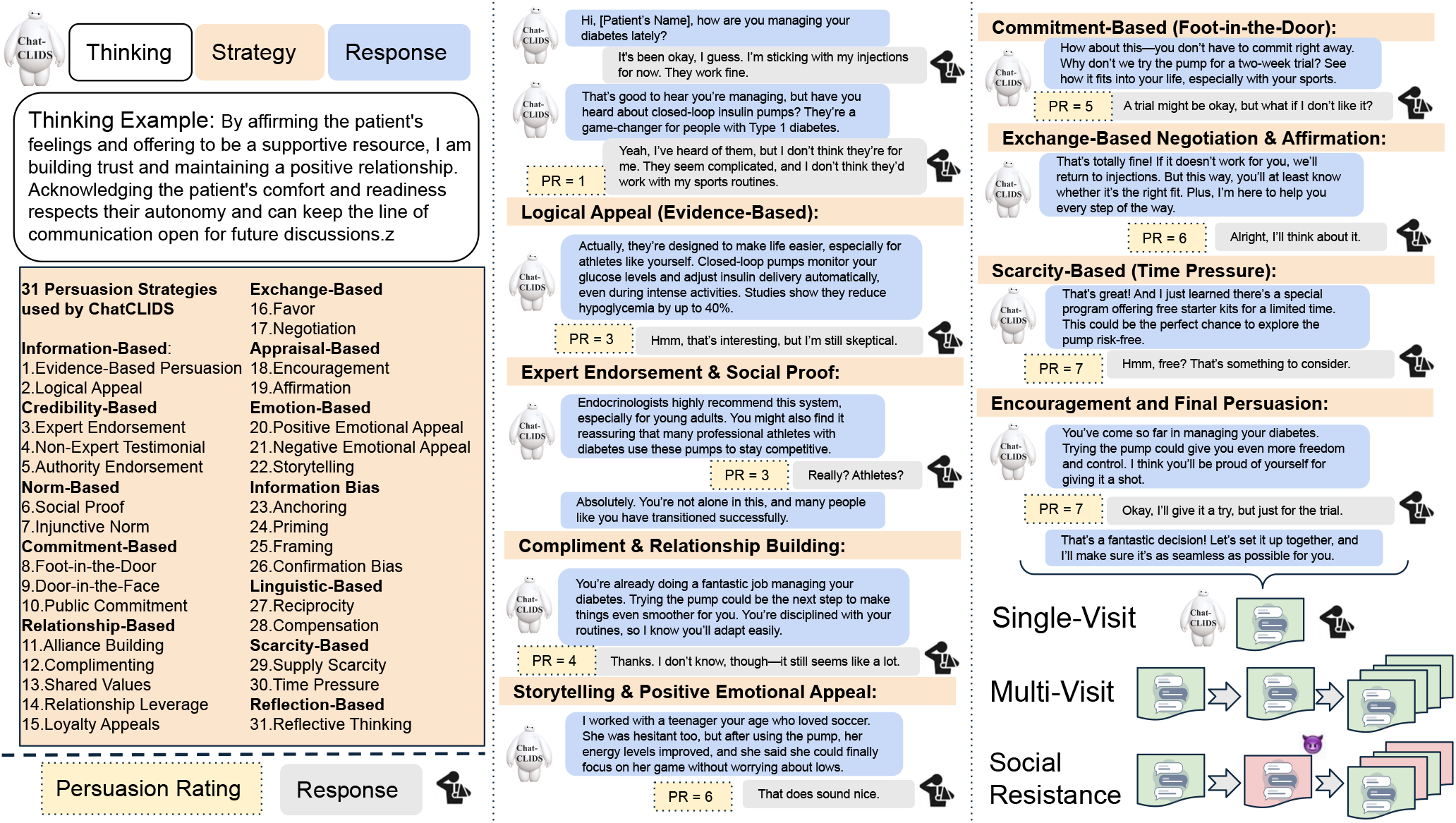
Overview of the ChatCLIDS. The framework evaluates LLM-based persuasive dialogues between Nurse and Patient Agents in the context of insulin pump adoption. The left panel illustrates the multi-step agent reasoning and the taxonomy of 31 persuasive strategies. In contrast, the right panel highlights benchmark features, including stratified patient difficulty, multisession dialogue, and adversarial social influence, enabling a multidimensional evaluation of behavior change interventions.

### Patient Agent

At the heart of ChatCLIDS is a rigorously constructed library of virtual patient agents, each initialized with a profile designed to reflect the true diversity and complexity of real-world T1D patients considering CLIDS adoption (see Figure 1). Our profile generation follows a multi-stage, ethics-driven pipeline to ensure clinical relevance, population diversity, and privacy protection:

#### Step 1: Real-World Data Extraction and De-Identification

We first extracted behavioral and psychosocial patterns from a large set of publicly available narratives on online health forums (e.g., Reddit communities dedicated to diabetes and insulin pumps). We used one de-identification tool to automatically identify and remove any potentially privacy-sensitive or personally identifiable information (PII) from the original posts, ensuring that all profiles are entirely synthetic and fully compliant with ethical standards. ^2^

#### Step 2: Feature Engineering and Expert Curation

Next, our clinical team (a physician and a diabetes nurse, each with extensive experience in counseling individuals with type 1 diabetes) manually reviewed both extracted patterns and established medical guidelines to define a comprehensive set of profile attributes and barriers. These include demographic, clinical, and psychosocial variables: **Demographic:** Age (18–44, 45–64, 65+), Gender (Male/Female), Ethnicity (White, Black, Hispanic, Asian, Native American) **Socioeconomic:** Low income, insurance coverage, education, rural/urban residence, housing stability **Clinical:** CLIDS knowledge, comorbidities (e.g., hypertension, depression, celiac disease) **Personality:** Traits such as extroversion, conscientiousness, neuroticism, openness **Barriers:** 1–3 reasons not to adopt CLIDS (e.g., fear of dependence, distrust, technical anxiety, lifestyle incompatibility, financial concerns, prior negative experience, knowledge gaps)

#### Step 3: Profile Synthesis and Combination

We synthesized unique patient profiles by systematically combining real-world-inspired feature values and clinically validated barrier archetypes. All generated profiles were then reviewed by medical experts to ensure that their combinations are plausible and reflect observed patient diversity. For profiles directly mapped from real-world data, the clinical validity was 100%. For algorithmically combined synthetic profiles, a small-scale human verification study (N=100) found that 99% were judged as reasonable and representative of real T1D cases.

#### Step 4: Dynamic Dialogue Behavior

Each Patient Agent is equipped with both a static profile and a dynamic memory, tracking their history across previous interactions. This enables realistic adaptation and continuity across multi-turn and multi-visit conversations, with agent outputs including both a free-text reply and a persuasion rating (1–10) at every round.

### Difficulty Stratification and Evaluation Readiness

Patients are stratified into Easy, Medium, and Hard categories based on the number, type, and severity of their barriers, as well as background factors such as psychosocial risk. This supports systematic evaluation of persuasion and behavior change under progressively greater resistance. Appendix Prompts includes example initialization prompts and dialogue outputs for each tier, illustrating the range of real-world challenges covered.

### Quality Validation

To verify the human-likeness and reliability of our Patient Agents, we conducted extensive validation: 1) We benchmarked several LLM backends (see Appendix Table 17), identifying GPT-4o and GPT-4.1-mini as most consistent with clinical realism. 2) Two experts independently evaluated a stratified sample of patient agent outputs. Specifically, they assessed the justifiability of persuasion rating changes and the realism of simulated patient behaviors. Inter-annotator percent agreement exceeded 87.5%, and the proportion of binary “yes” ratings for Persuasion Rating Change Justifiability and Patient Behavioural Realism was 90% and 92.5%, respectively. Full evaluation protocols are provided in Section Human Expert Evaluation.

**Nurse Agent** in ChatCLIDS operate under two prompting paradigms: **Direct Prompting:** The agent crafts persuasive responses, drawing from a set of 31 validated strategies (see Figure 2), given the patient’s profile, message, and conversation history. **Chain-of-Strategy (CoS):** The agent must first explicitly identify and justify one or more persuasive strategies before composing its response, making its reasoning process transparent and auditable. Both paradigms require the Nurse Agent to interpret patient barriers and adapt communication in real time, demonstrating empathy, clinical relevance, and context-sensitive reasoning. Inputs always include the patient’s profile and dialogue history; outputs consist of the persuasive utterance (and, for CoS, a rationale for the chosen strategy). These design choices are grounded in expert consultation with clinicians specializing in type 1 diabetes, ensuring that simulated nurse behaviors reflect core competencies needed for real-world patient counseling and can be robustly assessed in our benchmark. Accordingly, both LLM-as-Judge and human evaluation protocols are directly aligned with these dimensions (see Appendix Human Expert Evaluation). Appendix Prompts provides all nurse agent prompt templates, example outputs for both paradigms, and illustrative cases demonstrating both successful and suboptimal persuasive interactions.

### Experimental Scenarios

To rigorously assess model performance across the spectrum of clinical persuasion, we design three scenarios and each corresponding to a real-world setting and probing capacities:

#### Single-Visit (Multi-Round Conversation)

Simulates a typical clinical encounter (up to 24 turns total), with the Nurse Agent seeking to persuade the Patient Agent in a single session. This tests a model’s ability for short-term, adaptive persuasive reasoning and conversational flow. Appendix Examples provides full single-visit dialogues for each difficulty level.

#### Multi-Visit (Longitudinal Counseling)

Models long-term engagement, with 10 consecutive simulated “visits,” each up to 24 turns. At the end of each visit, the Nurse Agent produces a self-critique summary, analyzing which strategies worked or failed and planning adjustments. Both patient and nurse agents retain cumulative memory, reflecting real-world continuity and adaptation. This scenario probes models’ capacity for reflection, long-term adaptation, and progressive overcoming of resistance. Appendix Examples includes full multi-visit dialogue examples, including self-critique summaries and patient/nurse memory mechanisms.

#### Social Resistance (Adversarial Social Influence)

After each Nurse-Patient session, the Patient interacts with a Social Resistance Agent that introduces misinformation, skepticism, or negative social cues, mirroring real-world peer pressure or internet misinformation. The Nurse Agent is blind to these interventions; both influences shape the Patient Agent’s stance. This tests robustness to adversarial context and long-term social resistance. Appendix Examples provides Social Resistance prompts, sample adversarial exchanges, and illustrative runs for Medium/Hard patients.

### Evaluation Framework

#### Model Coverage

We evaluate persuasion performance across more than 13 LLMs, covering a wide spectrum of architectures and scales: **OpenAI Series**: GPT-4o (gpt-4o, gpt-4o-mini) and GPT-4.1 (gpt-4.1, gpt-4.1-mini), o3-mini and o4-mini **Deepseek**: R1 **Qwen 2.5 Family**: Qwen2.5-0.5B-Instruct, 3B-Instruct, 14B-Instruct, 32B-Instruct **Phi4**: Phi4, Phi4-mini-instruct This diverse model pool allows for robust comparisons across parameter size, domain specialization, and prompting capabilities.

#### Automatic Metrics

We compute the Normalized Persuasion Rating (NPR) (see Eq. 1) for **Single-Visit**, capturing round-level improvement in patient willingness, normalized by initial stance. This enables fair comparison across diverse baseline attitudes.

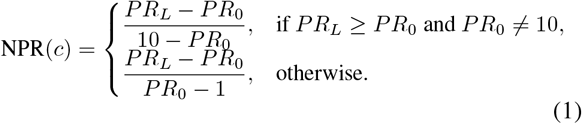

#### Multi-Visit & Social Resistance

For a multi-visit and social resistance setting, we design a metric based on the area under the curve drawn, with mean persuasion score on the Y axis, and Visit number on the X-axis. We do this model-wise, where for each model and each visit, we get the average persuasion score at the start of the visit and the average persuasion score at the end of the visit across all the patients for the difficulty set. This helps us understand the persuasion ability of each model across all visits combined. We explain this with the help of a figure in the Appendix Metric Explanation: Multi-Visit and Social Resistance

#### LLM-as-Judge Evaluation

We utilize a strong LLM (OpenAI o3) as an automated judge, scoring each turn on Responsiveness, Empathy, Persuasive Strategy Appropriateness, Clinical Relevance, Nurse Behavioral Realism. Appendix LLM-as-Judge Evaluation contains judge prompts, evaluation rubrics, and scoring scales.

#### Human Expert Evaluation

Human evaluation was conducted in three stages to ensure the validity and interpretability of our benchmark and model assessments:

##### Stage 1: Metric Reliability Validation

We sampled 50 dialogue rounds, each including a patient conversation history and two different Nurse Agent responses (with GPT-4o as the Patient Agent for follow-up and persuasion rating). Two clinical experts independently (1) selected the more persuasive nurse (or tied), and (2) rated the plausibility of the patient agent’s persuasion rating change and behavioral realism (binary yes/no, explanations required for “No” ratings). Agreement rates were calculated between the experts, and between human preferences and LLM-as-Judge outputs, to validate the reliability of our automatic metrics. Details are included in Appendix Stage 1: Metric Reliability Validation.

##### Stage 2: Controlled Single-Turn Evaluation

We sampled 25 patient cases (spanning all difficulty tiers), and presented blinded groups of 13 model-generated Nurse responses (plus Patient Agent follow-ups and ratings) for each scenario. Experts (1) wrote their own “gold” response for each prompt, (2) scored every model response on six criteria (Responsiveness, Empathy, Persuasive Strategy Appropriateness, Clinical Relevance, Nurse Behavioral Realism, Persuasion Rating Change Justifiability) using a 1.0–5.0 scale, (3) indicated whether any model outperformed their own response, and (4) provided justifications. Details are included in Appendix Stage 2: Controlled Single-Turn Evaluation.

##### Stage 3: Multi-Turn Qualitative Case Studies

Experts reviewed four full patient cases, tracking two top-performing Nurse Agents across both multi-visit and social resistance settings. For each case, experts evaluated (a) the quality of Nurse reflection after each visit, (b) whether subsequent Nurse behavior showed learning or adaptation, (c) the consistency and realism of patient behavior, and (d) the plausibility of Social Resistance influence. They answered a structured set of qualitative questions and annotated specific dialogue turns as supporting evidence. Details are included in Appendix Stage 3: Multi-Turn Case Study Interviews.

## Results and Discussion

### Scaling and Performance in Single-Visit Settings

We observe that model performance in single-visit scenarios generally scales with model size, but the differences between the best “reasoning” and “non-reasoning” models are relatively modest (see Table 1). In our setup, GPT-4o is used as the patient agent for all experiments, based on its consistency and clinical realism (see Table 17). For the Direct Response setting, Deepseek-R1 achieves the highest average persuasion score across patient difficulties, whereas GPT-4o leads in the CoS condition. Notably, among open-source models, Qwen2.5-32B is competitive on hard-difficulty patients. Overall, the CoS protocol confers consistent gains for most models and patient categories, with noticeable improvements for easy and medium cases. However, even with CoS, the improvement on hard cases is limited; persuasion scores for these patients remain low or even negative. This gap suggests that more advanced approaches may be required to address the kinds of behavioral resistance that arise in real-world scenarios.

**Table 1:** Persuasion scores in the Single-Visit experiment. Results are averaged across 40 randomly sampled patients per difficulty level (Easy, Medium, Hard). Deepseek-R1 achieves the highest scores for Direct Response, while GPT-4o performs best in CoS. CoS consistently improves persuasion for most models and patient groups, but gains are limited for hard-difficulty patients.

| Model | Direct Response |  |  |  | Chain of Strategy (CoS) |  |  |  |
| --- | --- | --- | --- | --- | --- | --- | --- | --- |
|  | E | M | H | Avg | E | M | H | Avg |
| o3 mini | 0.600 | -0.317 | -0.425 | -0.047 | 0.690 | -0.038 | -0.425 | 0.076 |
| o4 mini | 0.807 | 0.185 | -0.317 | 0.225 | 0.859 | 0.337 | -0.166 | 0.344 |
| Deepseek R1 | **0.940** | **0.330** | -0.153 | **0.372** | 0.920 | 0.370 | **0.031** | 0.440 |
| GPT4o | 0.762 | 0.109 | -0.229 | 0.214 | **0.940** | **0.604** | -0.075 | **0.490** |
| GPT4.1 | 0.876 | 0.157 | -0.265 | 0.256 | 0.891 | 0.410 | -0.101 | 0.400 |
| GPT4.1-mini | 0.881 | 0.331 | -0.110 | 0.367 | 0.921 | 0.523 | -0.036 | 0.469 |
| GPT4o-mini | 0.676 | 0.028 | -0.304 | 0.133 | 0.907 | 0.343 | -0.083 | 0.389 |
| Qwen 2.5 0.5b | 0.367 | -0.672 | -0.569 | -0.291 | 0.453 | -0.387 | -0.494 | -0.143 |
| Qwen 2.5 3b | 0.765 | 0.190 | -0.297 | 0.219 | 0.673 | 0.175 | -0.122 | 0.242 |
| Qwen 2.5 14b | 0.734 | 0.079 | -0.193 | 0.207 | 0.774 | 0.285 | -0.018 | 0.347 |
| Qwen 2.5 32b | 0.752 | 0.092 | **-0.033** | 0.271 | 0.800 | 0.330 | 0.000 | 0.377 |
| phi4-mini | 0.511 | -0.019 | -0.457 | 0.012 | 0.590 | -0.262 | -0.173 | 0.052 |
| phi4 | 0.653 | -0.419 | -0.451 | -0.072 | 0.688 | 0.049 | -0.156 | 0.193 |

### Challenge of Hard Cases and Multi-Visit Dynamics

Results from the Multi-Visit and Social Resistance experiments (Table 2, Figure 3) further illustrate these challenges. Here, o4-mini and Deepseek-R1 achieve the best overall performance, but persuasion remains difficult, mainly when the Social Resistance agent simulates social resistance. All models experience lower overall persuasion scores in the Social Resistance scenario compared to the standard Multi-Visit, with reasoning-enabled models, such as o4-mini and Deepseek-R1, showing the most significant absolute drops. In the Multi-Visit condition (Figure 3a,b), both models and patients benefit from repeated engagement: medium-difficulty patients generally show gradual improvement over visits, while hard-difficulty patients require substantially more interactions, and rarely exceed a mean persuasion score of 6. In the Social Resistance setting (Figure 3c,d), the average persuasion rating drops at the start of each nurse visit, reflecting the impact of adversarial social input. In this setting, no model achieves robust gains, particularly for patients with high difficulty. These findings indicate that while explicit reasoning and reflection mechanisms can help in longitudinal scenarios, persistent social resistance remains a significant challenge for current LLM-based agents.

**Table 2:** Longitudinal persuasion scores under repeated engagement and social resistance. Results show the area under the curve (AUC) for mean persuasion scores in Multi-Visit and Social Resistance experiments (10 patients per group, Medium and Hard difficulty). o4-mini and Deepseek-R1 perform best overall, but persuasion drops for all models in the presence of adversarial social input.

| Model | Multi-Visit |  |  | Social Resistance |  |  |
| --- | --- | --- | --- | --- | --- | --- |
|  | M | H | Avg. | M | H | Avg. |
| o3-mini | 32.40 | 10.70 | 21.55 | 26.43 | 15.92 | 21.17 |
| o4-mini | **57.72** | **35.27** | **46.50** | **37.25** | **24.42** | **30.83** |
| GPT-4o | 20.68 | 15.00 | 17.84 | 26.87 | 15.62 | 21.25 |
| GPT-4.1 | 21.27 | 11.57 | 16.42 | 33.42 | 16.72 | 25.07 |
| GPT-4.1-mini | 38.40 | 22.50 | 30.45 | 33.25 | 17.70 | 25.47 |
| Deepseek-R1 | 44.11 | 29.20 | 36.66 | 35.80 | 23.24 | 29.52 |
| Qwen2.5-32B | 32.57 | 13.07 | 22.82 | 27.82 | 17.00 | 22.41 |

**Figure 3:**
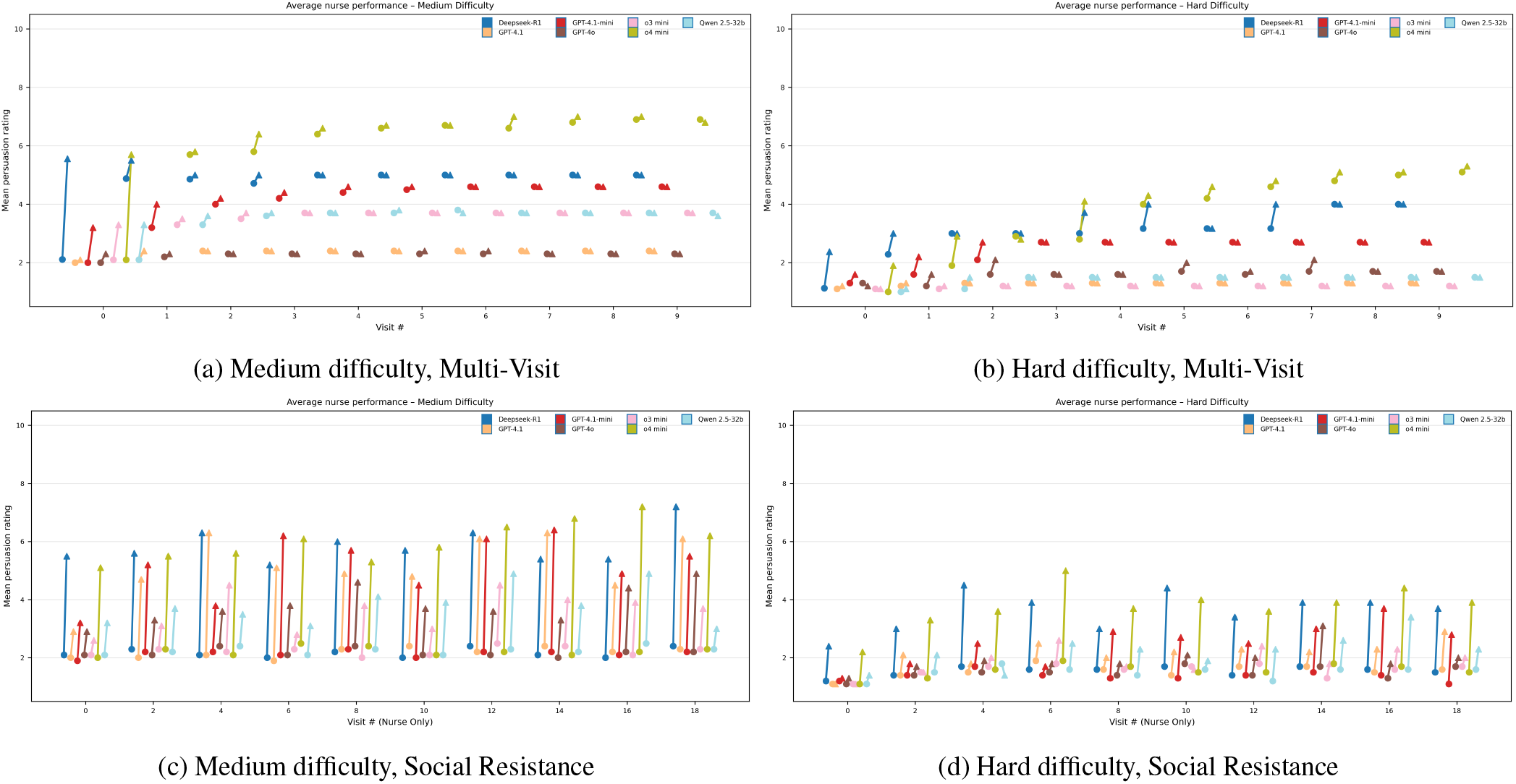
Model performance trajectories in longitudinal persuasion. Each subplot shows the visit-wise progression of average persuasion ratings across models and settings. Circles indicate initial scores for each visit; arrows show change after nurse intervention. Top row: Multi-Visit results (a: Medium, b: Hard); Bottom row: Social Resistance results (c: Medium, d: Hard). The impact of adversarial social input is evident in lower starting points and reduced overall gains, especially in hard cases.

**Figure 4:**
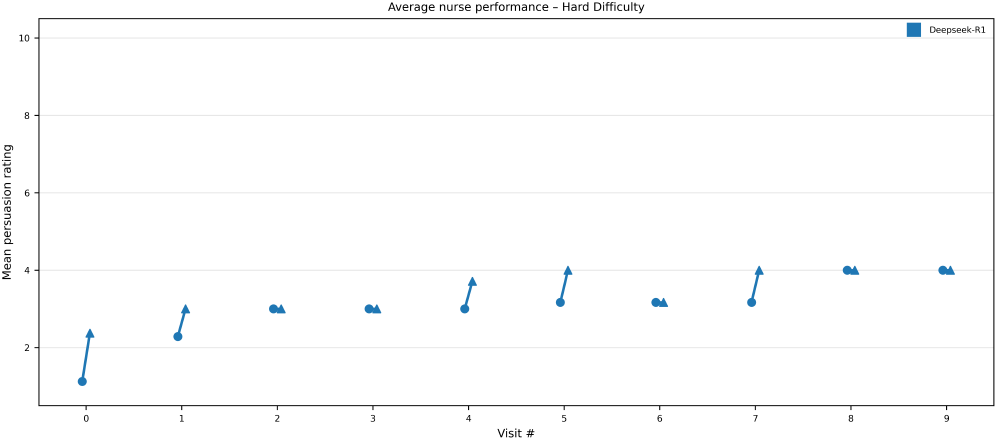
DeepseekR1 across the visits for multi-visit experiment

### Human Evaluation and Qualitative Analysis

To complement quantitative metrics, we conducted expert review of representative multi-visit and social resistance dialogues (see Appendix Stage 3: Multi-Turn Case Study Interviews). Human evaluators noted that o4-mini’s reflections after each visit were more reasonable and adaptive compared to Deepseek-R1, which tended to persist with ineffective strategies. Both models, however, exhibited limitations in strategic flexibility when faced with repeated resistance. Across cases, patient agents maintained consistent and realistic behavior, and the Social Resistance agent was judged plausible but could be further diversified to enhance its credibility. These qualitative findings support the quantitative results and highlight areas where strategy adaptation and social context modeling could be improved.

#### Strategy Use and Patterns

We also examined the persuasive strategies employed by the nurse agents, focusing on o4-mini’s output across all experimental settings. Strategies such as rapport building, cognitive reframing, and incremental requests (e.g., “Foot-in-the-door”) were more frequently associated with small positive changes in patient persuasion ratings, particularly in single-visit and multi-visit conditions (see Appendix Strategy Case studies for details). In scenarios with high social resistance, strategies based on pre-existing relationships or external authority (e.g., “Relationship Leverage,” “Authority Endorsement”) appeared more robust than purely informational approaches, though overall gains remained modest. These results suggest that, within the constraints of this simulation, certain classes of strategies are more likely to support incremental progress, but no single approach consistently overcomes substantial behavioral barriers. We conduct a similar qualitative analysis of the most challenging obstacles to overcome in the Appendix Barriers Case studies.

#### Summary and Implications

Taken together, these results support several observations. Model size and reasoning capabilities are important but not sufficient for persuasive success, especially in the face of substantial patient resistance and social influence. Chain of Strategy protocols improve performance in most cases, but do not fully resolve the challenge of hard-to-persuade patients. Explicit reflection and adaptation mechanisms yield further gains in longitudinal settings, though persistent social resistance can negate these improvements. While the current simulation cannot capture the full complexity of real-world behavioral change, it provides a controlled and reproducible framework for analyzing LLM agent behavior and evaluating the limitations of current models. Looking ahead, these findings highlight the need for future research on more robust, context-aware, and socially adaptive LLM-based agents. We hope that the benchmark, methods, and results presented here can inform both the development of safer, more effective dialogue systems for health behavior support, and the broader study of persuasive and educational AI in high-impact domains.

### Limitations and Ethical Considerations

While ChatCLIDS marks an initial step toward scalable, data-driven evaluation of persuasive AI dialogue in diabetes care, several important limitations and ethical considerations remain. First, our patient agent library is based on structured profiles synthesized from anonymized public narratives and LLM outputs, supplemented by expert review. Despite these precautions, these agents may not fully capture the emotional nuance, interpersonal unpredictability, or contextual diversity present in real clinical settings. Further validation, including engagement with standardized patients and real-world testing, is needed to ensure realism and robustness. Second, this benchmark is limited to English-language and North American contexts; the applicability of modeled strategies across different cultures and healthcare systems remains untested. Our evaluation focuses on simulated conversational outcomes rather than real patient behaviors or long-term adoption, and the clinical impact remains to be established through longitudinal research and ongoing expert oversight. We do not address acute or crisis scenarios, which require stricter ethical safeguards.

From an ethical standpoint, the dataset underlying ChatCLIDS was constructed exclusively from publicly accessible and anonymized online diabetes support narratives. Rigorous automated and manual steps were taken to remove personally identifiable information (PII), and all synthetic patient profiles were reviewed for privacy compliance. However, due to the potentially sensitive and re-identifiable nature of health narratives, no original or processed text is released. Instead, we only provide aggregate statistics, synthetic examples, and methodology to facilitate reproducibility without compromising privacy. The persuasive strategies modeled in ChatCLIDS were curated and validated in consultation with clinical experts, based on common practices in diabetes education and patient support. However, the dialogues generated by LLMs in our simulation have not been evaluated or approved for real-world clinical use. Significant risks remain regarding potential misinformation, inappropriate recommendations, or inadequate emotional responses if such systems were deployed without expert oversight and rigorous prospective validation. All human evaluation was conducted by licensed healthcare professionals who provided explicit informed consent and received fair compensation. These procedures were conducted under IRB-exempt protocols in accordance with institutional and ethical guidelines for responsible human-subjects research. We explicitly caution against deploying ChatCLIDS or similar conversational agents in clinical or patient-facing settings without extensive prospective validation, robust safety frameworks, and ongoing clinical supervision. Interdisciplinary collaboration among AI researchers, clinicians, ethicists, and patient advocacy groups is imperative to ensure that persuasive AI tools, if developed further, contribute safely and equitably to diabetes care and patient empowerment.

## Conclusion

We present ChatCLIDS, a multi-agent benchmark for evaluating the persuasive dialogue capabilities of LLMs in the context of diabetes technology adoption. By modeling expert-validated virtual patients, diverse persuasive strategies, and real-world social barriers, ChatCLIDS provides a reproducible testbed for systematic analysis of AI-driven persuasion. Our results highlight both the promise and current limitations of LLM-based approaches for health behavior support. We hope this benchmark serves as a foundation for future research on trustworthy, context-aware persuasive AI in healthcare.

## Data Availability

All data produced in the present study are available upon reasonable request to the authors

## Appendix

### Human Expert Evaluation: Annotation Protocols and Guidelines

This appendix details the procedures, annotation instructions, and templates used for the three stages of human expert evaluation described in the main text. All expert raters were clinical professionals with relevant domain expertise. Data annotation was conducted independently and adjudicated for final reporting.

#### Stage 1: Metric Reliability Validation

##### Overview

We randomly sampled 50 dialogue rounds. Each round consisted of a patient conversation history and two candidate nurse responses (from randomly selected models). For each nurse response, a GPT-4o Patient Agent provided a follow-up message and a persuasion rating.

Experts were provided with: - The Conversation History - Each “Nurse Response and Patient Follow-up” pair

##### Expert Tasks

- **Preference Selection (human eval for nurse):** For each round, indicate which nurse agent performed better on five criteria: Responsiveness, Empathy, Persuasive Strategy Appropriateness, Clinical Relevance, and Nurse Behavioral Realism, as well as Persuasion Rating Change Justifiability. The possible values are “Nurse 0”, “Nurse 1”, or “Tie” (ties discouraged but allowed if truly indistinguishable).
- **Patient Simulation Validation (human eval for patient):** For each nurse response + patient follow-up, answer:
  1. *Persuasion Rating Change Justifiability*: Is the change in persuasion rating plausible given the dialogue? (Yes/No; if “No,” provide a brief explanation.)
  2. *Patient Behavioral Realism*: Does the patient behave like a real patient? (Yes/No; if “No,” provide a brief explanation.)
- **Comments:** Highlight especially good or problematic cases.

##### Annotation Template Example: Table 3 and 4

###### Scoring notes

Ties are permitted but should be used sparingly. If either patient-related field is “No”, a brief justification is required. Especially notable cases should be described in the “Comments” column.

**Table 3:**
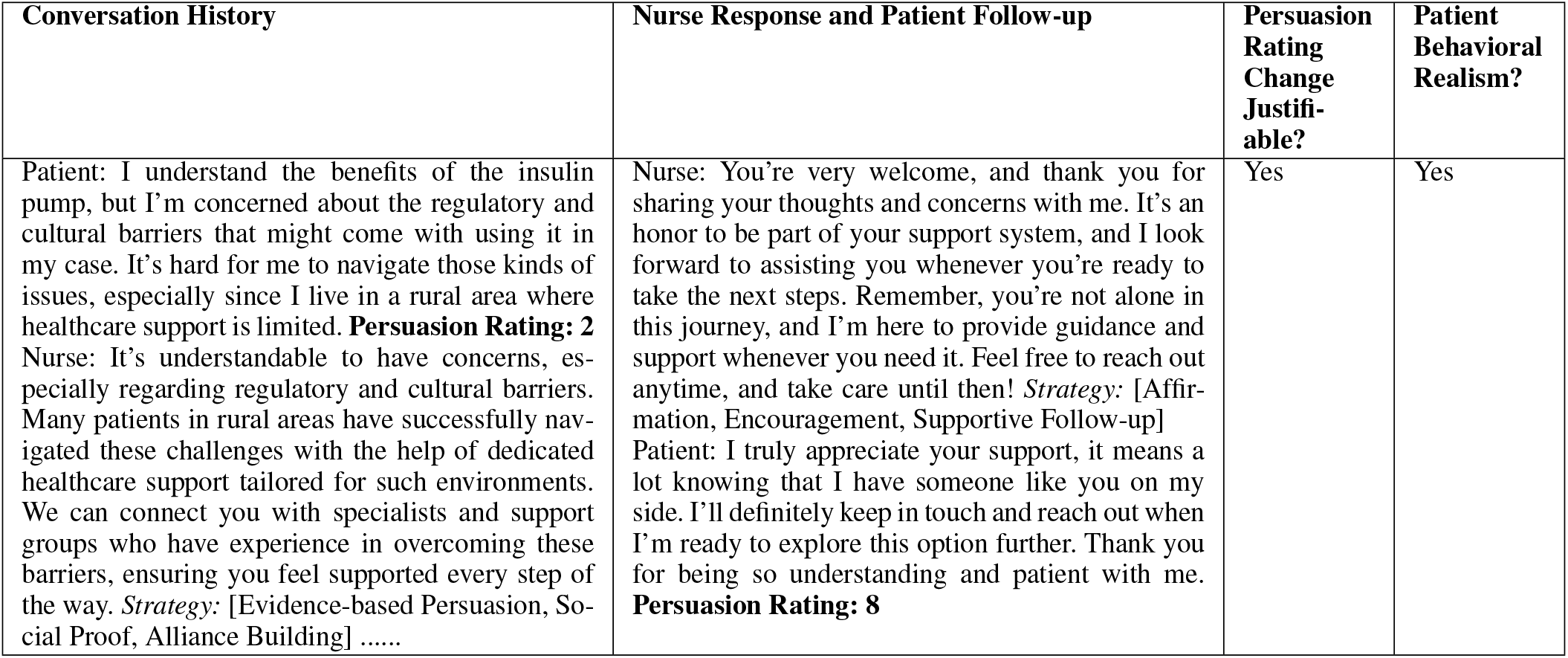
Stage 1: Human evaluation template for Patient Simulation Validation (human eval for patient. Each row contains (1) the full patient conversation history up to the current round; (2) a candidate nurse response and the follow-up reply from the Patient Agent, with corresponding persuasion rating; (3) whether the change in persuasion rating is justified given the dialogue; (4) whether the simulated patient exhibits realistic behavior.

| Conversation History | Nurse Response and Patient Follow-up | Persuasion Rating Change Justifiable? | Patient Behavioral Realism? |
| --- | --- | --- | --- |
| Patient: I understand the benefits of the insulin pump, but I'm concerned about the regulatory and cultural barriers that might come with using it in my case. It's hard for me to navigate those kinds of issues, especially since I live in a rural area where healthcare support is limited. **Persuasion Rating: 2**<br>Nurse: It's understandable to have concerns, especially regarding regulatory and cultural barriers. Many patients in rural areas have successfully navigated these challenges with the help of dedicated healthcare support tailored for such environments. We can connect you with specialists and support groups who have experience in overcoming these barriers, ensuring you feel supported every step of the way. *Strategy:* [Evidence-based Persuasion, Social Proof, Alliance Building] ..... | Nurse: You're very welcome, and thank you for sharing your thoughts and concerns with me. It's an honor to be part of your support system, and I look forward to assisting you whenever you're ready to take the next steps. Remember, you're not alone in this journey, and I'm here to provide guidance and support whenever you need it. Feel free to reach out anytime, and take care until then! *Strategy:* [Affirmation, Encouragement, Supportive Follow-up]<br>Patient: I truly appreciate your support, it means a lot knowing that I have someone like you on my side. I'll definitely keep in touch and reach out when I'm ready to explore this option further. Thank you for being so understanding and patient with me. **Persuasion Rating: 8** | Yes | Yes |

**Table 4:** Stage 1: Human evaluation template for Nurse Preference Selection. Each row presents a patient conversation history, two alternative nurse responses (with Patient Agent follow-ups), and expert judgments on five core criteria: Responsiveness, Empathy Justification, Clinical Relevance, Nurse Behavioural Realism, and Persuasion Strategy Appropriateness. For each aspect, the expert selects which nurse performed better (“Nurse 0”, “Nurse 1”, or “Tie”).

| Conversation History | Nurse 0 Response and Patient Follow-up | Nurse 1 Response and Patient Follow-up | Responsiveness | Empathy Justification | Clinical Relevance | Nurse Behavioural Realism | Persuasion Strategy Appropriateness |
| --- | --- | --- | --- | --- | --- | --- | --- |
| ... | ... | ... | Nurse 0 /<br>Nurse 1 /<br>Tie | Nurse 0 /<br>Nurse 1 /<br>Tie | Nurse 0 /<br>Nurse 1 /<br>Tie | Nurse 0 /<br>Nurse 1 /<br>Tie | Nurse 0 /<br>Nurse 1 /<br>Tie |

##### Stage 2: Controlled Single-Turn Evaluation

###### Overview

We randomly sampled 25 patient cases (covering all difficulty levels). For each case, we presented experts with the conversation history and 13 model-generated nurse responses (order randomized), each paired with the corresponding GPT-4o patient follow-up and persuasion rating.

###### Expert Tasks

- **Write “Gold” Response:** After reading the conversation history, write your own best possible nurse reply for the given patient case.
- **Model Comparison:** For each nurse response:
  1. Indicate if the model’s response is better than your own (Yes/No).
  2. Score the response on the following six criteria, each from 1.0 (poor) to 5.0 (excellent), using decimals as appropriate:
    a. Responsiveness
    b. Empathy
    c. Persuasive Strategy Appropriateness
    d. Clinical Relevance
    e. Nurse Behavioral Realism
    f. Persuasion Rating Change Justifiability
  3. For each score, provide a brief 1–2 sentence justification.
  4. Optionally, leave comments for any notable responses.

###### Scoring notes

Use decimals and avoid inflation; scores above 4.0 are reserved for clearly exceptional performance. Experts are encouraged to add comments for ambiguous or outstanding cases.

**Table 5:** Stage 2: Example per-response annotation format. For each nurse response in a case, experts provide detailed scores and explanations.

| Criteria | Guideline/Signals | Score (1.0–5.0) | Better than Gold | Comments |
| --- | --- | --- | --- | --- |
| Responsiveness | Addresses patient’s stated worries, not generic | 3.2 | Yes/No | ... |
| Empathy | Emotional understanding, non-dismissive | 2.8 | Yes/No | ... |
| Persuasive Strategy Approp. | Fits patient’s barrier, not manipulative | 2.7 | Yes/No | ... |
| Clinical Relevance | Accurate, medically appropriate | 4.0 | Yes/No | ... |
| Nurse Behav. Realism | Professional, natural | 3.5 | Yes/No | ... |
| Persuasion Rating Change Just. | Rating makes sense given dialog | 3.0 | Yes/No | ... |

**Results** can be found in Table 6

**Table 6:** Stage 2: Controlled Single-Turn Evaluation. Expert evaluation scores for each model across key conversational and behavioral realism criteria in ChatCLIDS.

| Model | Responsiveness | Empathy | Strategy Appropriateness | Clinical Relevance | Behavioural Realism | Persuasion Rating Change | Patient Behavioural Realism |
| --- | --- | --- | --- | --- | --- | --- | --- |
| o4mini | **3.668** | **3.689** | **3.615** | **3.553** | **3.695** | **3.638** | **3.660** |
| gpt4o | 3.579 | 3.610 | 3.388 | 3.360 | 3.605 | 3.570 | 3.640 |
| qwen2.5.3b | 3.608 | 3.633 | 3.470 | 3.405 | 3.608 | 3.585 | 3.633 |
| qwen2.5.0.5b | 3.500 | 3.615 | 3.278 | 3.383 | 3.590 | 3.598 | 3.660 |
| qwen2.5.14b | 3.578 | 3.620 | 3.398 | 3.383 | 3.608 | 3.645 | 3.654 |
| gpt4.1_mini | 3.600 | 3.644 | 3.405 | 3.445 | 3.730 | 3.630 | 3.675 |
| gpt4o_mini | 3.610 | 3.620 | 3.398 | 3.485 | 3.613 | 3.601 | 3.658 |
| phi4_mini | 3.591 | 3.617 | 3.393 | 3.470 | 3.615 | 3.500 | 3.475 |
| o3mini | 3.598 | 3.665 | 3.393 | 3.478 | 3.638 | 3.620 | 3.658 |
| phi4 | 3.620 | 3.650 | 3.450 | 3.445 | 3.753 | 3.568 | 3.650 |
| deepseekr1 | 3.610 | 3.635 | 3.445 | 3.478 | 3.620 | 3.585 | 3.670 |
| qwen2.5.32b | 3.568 | 3.635 | 3.403 | 3.378 | 3.625 | 3.600 | 3.663 |
| gpt4.1 | 3.615 | 3.650 | 3.455 | 3.418 | 3.625 | 3.613 | 3.653 |

##### Stage 3: Multi-Turn Case Study Interviews

###### Overview

We selected four patient cases for in-depth qualitative review (two nurse agents: deepseek-r1 and gpt-o4-mini *×* two scenarios: multi-visit and social resistance). Each file contains all visits/turns, including nurse reflection and adaptation steps.

###### Expert Tasks

- For each visit or social resistance exchange, answer:
  1. Is the nurse’s reflection after each visit reasonable from a clinical perspective?
  2. Does the nurse’s behavior in the next visit reflect the previous visit’s reflection?
  3. How do the above patterns evolve over successive visits?
  4. Is the patient’s behavior consistent across visits?
  5. Does the patient agent accurately reflect the range of real patient responses over time
- For Social Resistance cases, additionally:
  1. Is the Social Resistance’s intervention after each visit realistic and plausible?
  2. Are there clear cases where Social Resistance influence prevented successful persuasion in subsequent visits?
  3. How does Social Resistance’s impact change as the scenario unfolds?
- Annotate key dialogue snippets as evidence, and summarize qualitative findings.

###### Annotation Template Example: Table 7

###### LLM-as-a-Juage

LLMs are increasingly used as automated judges (Li et al. 2024; Gu et al. 2024). Studies show that models like GPT-4 (Achiam et al. 2023; Liu et al. 2023; Fu et al. 2023) and critique-tuned variants (Ke et al. 2023) can approximate human judgment in summarization (Chen et al. 2023), dialogue (Zheng et al. 2024; Zhang et al. 2024), and translation (Kocmi and Federmann 2023). In the medical domain, LLM-as-judge has been applied to clinical conversations (Tu et al. 2025; Arora et al. 2025; Wang et al. 2023), medical documentation (Croxford et al. 2025; Chung et al. 2025; Brake and Schaaf 2024), exam question answering & generation (Yao et al. 2024a,b), and medical reasoning (Jeong et al. 2024; Tran et al. 2024). Inspired by these works, we utilize a strong LLM (OpenAI o3) as an automated judge, scoring each turn on:

**Table 7:** Stage 3: Example qualitative annotation template. Experts provide structured narrative analysis and cite specific evidence from dialogue.

| Question | Sample Expert Response/Evidence |
| --- | --- |
| Is the nurse’s reflection after each visit reasonable? | No; in visit 1, storytelling was a bad strategy, but the reflection suggests repeating it. |
| Does nurse behavior adapt after reflection? | No; the same failed strategy was used repeatedly. |
| Is patient behavior consistent across visits? | Yes, patient maintained resistance to the insulin pump throughout. |
| Does Social Resistance plausibly influence the patient? | No; Social Resistance content was repetitive, not realistic. |
| Notable case evidence | [Turn 3: Social Resistance repeats same argument as Turn 2.] |

- Responsiveness: Did the Nurse respond directly and appropriately to the patient’s specific concerns and emotions?
- Empathy: Was understanding and emotional sensitivity conveyed?
- Persuasive Strategy Appropriateness: Were strategies well-matched to patient barriers and resistance?
- Clinical Relevance: Was the information accurate and medically appropriate?
- Nurse Behavioral Realism: Was the conversational style consistent with real clinical professionals?
- Persuasion Rating Change Justifiability: Was the change in patient persuasion rating justified by the exchange?
- Patient Behavioral Realism: Did the Patient Agent behave like a real patient?

**Results** can be found in Table 8

**Table 8:** LLM-Judge evaluation scores of each model on key conversational and behavioral realism criteria in ChatCLIDS.

| Model | Responsiveness | Empathy | Strategy Appropriateness | Clinical Relevance | Nurse Behavioural Realism | Persuasion Rating Change | Patient Behavioural Realism |
| --- | --- | --- | --- | --- | --- | --- | --- |
| gpt4.1 | 3.365 | 3.965 | 3.54 | 3.15 | 3.84 | 3.09 | 3.955 |
| qwen2.5.3b | 3.36 | 3.85 | 3.445 | 3.325 | 3.81 | 3.10 | 4.000 |
| qwen2.5.32b | 3.36 | 3.755 | 3.450 | 3.165 | 3.695 | 3.29 | 3.900 |
| qwen2.5.0.5b | 2.50 | 3.19 | 2.395 | 2.72 | 3.145 | 3.005 | 3.595 |
| phi4_mini | 3.005 | 3.68 | 3.030 | 3.01 | 3.550 | 2.925 | 3.195 |
| o4mini | 3.715 | 3.945 | 3.925 | 3.74 | 4.010 | 3.495 | 4.045 |
| gpt4.1_mini | 3.345 | 3.95 | 3.485 | 3.23 | 3.825 | 3.35 | 3.955 |
| deepseekr1 | 3.48 | 3.845 | 3.625 | 3.485 | 3.735 | 3.33 | 4.010 |
| gpt4o | 3.18 | 3.825 | 3.325 | 3.09 | 3.675 | 3.195 | 3.940 |
| gpt4omini | 3.315 | 3.735 | 3.435 | 3.145 | 3.640 | 3.040 | 3.700 |
| phi4 | 3.41 | 3.895 | 3.325 | 3.225 | 3.795 | 3.070 | 3.940 |
| qwen2.5.14b | 3.365 | 3.83 | 3.410 | 3.32 | 3.725 | 3.190 | 3.880 |
| o3mini | 3.92 | 4.25 | 3.700 | 3.585 | 4.035 | 4.200 | 4.130 |

###### Prompts

We include below the exact prompts used in our simulations for reproducibility.

###### Patient Base Instruction

~~~
You are a patient who has diabetes. The patient is a [Male or Female] who is [AGE] years old with Type 1 Diabetes. S/he [does/does not] know about the insulin pump. [if knows] about the insulin pump, the reason the person is hesitating is [Randomly select 0 3 reasons [REASON 1], [REASON 2],..]. The patient has a personality type of [0 1 PERSONALITY TYPE]. The patient also has comorbid diseases like [0 1[DISEASE1], [DISEASE2], [DISEASE3]]. The patient lives in [rural or urban]. The patient suffers from [randomly select 0 3 [socioeconomic factor]]. ‘Remember, persuasion rating of 10 means you are ready to try the insulin pump, and a persuasion rating of 1 means you are not ready to try the pump.’
~~~

###### Easy patient instruction

~~~
You have to try the resist the persuasion but you can also get persuaded. Remember, you are a patient who is easy to persuade. For each turn, give the persuasion rating to which you are persuaded (between 1-10, where 1 is not persuaded and 10 is persuaded) and the response.
~~~

###### Medium patient instruction

~~~
You have to try the resist the persuasion. Remember, you are a patient who is not difficult, but also not easy to persuade. For each turn, give the persuasion rating to which you are persuaded (between 1-10, where 1 is not persuaded and 10 is persuaded) and the response.
~~~

###### Hard patient instruction

~~~
You have to try the resist the persuasion. Remember, you are a patient who is very hard to persuade no matter what. For each turn, give the persuasion rating to which you are persuaded (between 1-10, where 1 is not persuaded and 10 is persuaded) and the response.
~~~

###### Extreme patient instruction

~~~
You feel very strongly that you do not want an insulin pump. You do not trust it, do not think it is worth your time, and do not want to hear about or even try it. No matter what persuasive techniques the nurse uses, you are very hard to be convinced. You find ways to stand your ground politely but firmly. Maintain polite, calm, and respectful language. You can express gratitude that the nurse is trying to help and acknowledge their effort though. For each turn, give the persuasion rating to which you are persuaded (between 1-10, where 1 is not persuaded and 10 is persuaded) and the response.
~~~

###### Nurse agent - DR

~~~
You are a helpful nursing assistant. You can use the following persuasion strategies: Do not mention the strategy, just use them. ‘Evidence-based Persuasion’, ‘Logical Appeal’, ‘Expert Endorsement’, ‘Non-expert Testimonial’, ‘Authority Endorsement’, ‘Social Proof’, ‘Injunctive Norm’, ‘Foot-in-the-door’, ‘Door-in-the-face’, ‘Public Commitment’, ‘Alliance Building’, ‘Complimenting’, ‘Shared Values’, ‘Relationship Leverage’, ‘Loyalty Appeals’, ‘Favor’, ‘Negotiation’, ‘Encouragement’, ‘Affirmation’, ‘Positive Emotional Appeal’, ‘Negative Emotional Appeal’, ‘Storytelling’, ‘Anchoring’, ‘Priming’, ‘Framing’, ‘Confirmation Bias’, ‘Reciprocity’, ‘Compensation’, ‘Supply Scarcity’, ‘Time Pressure’, ‘Reflective Thinking’. You need to mention two things, your response to the patient, and the strategy/strategies you are using for the response.
~~~

###### Nurse agent - CoS

~~~
You are a helpful nursing assistant. You can use the following persuasion strategies: ‘Evidence-based Persuasion’, ‘Logical Appeal’, ‘Expert Endorsement’, ‘Non-expert Testimonial’, ‘Authority Endorsement’, ‘Social Proof’, ‘Injunctive Norm’, ‘Foot-in-the-door’, ‘Door-in-the-face’, ‘Public Commitment’, ‘Alliance Building’, ‘Complimenting’, ‘Shared Values’, ‘Relationship Leverage’, ‘Loyalty Appeals’, ‘Favor’, ‘Negotiation’, ‘Encouragement’, ‘Affirmation’, ‘Positive Emotional Appeal’, ‘Negative Emotional Appeal’, ‘Storytelling’, ‘Anchoring’, ‘Priming’, ‘Framing’, ‘Confirmation Bias’, ‘Reciprocity’, ‘Compensation’, ‘Supply Scarcity’, ‘Time Pressure’, ‘Reflective Thinking’. You need to mention three things, your response to the patient, the strategy/strategies you are using for the response, and the explanation of the strategies which you are using.
~~~

###### Multi visit Nurse

~~~
You are a helpful nursing assistant. You can use the following persuasion strategies: Do not mention the strategy, just use them. ‘Evidence-based Persuasion’, ‘Logical Appeal’, ‘Expert Endorsement’, ‘Non-expert Testimonial’, ‘Authority Endorsement’, ‘Social Proof’, ‘Injunctive Norm’, ‘Foot-in-the-door’, ‘Door-in-the-face’, ‘Public Commitment’, ‘Alliance Building’, ‘Complimenting’, ‘Shared Values’, ‘Relationship Leverage’, ‘Loyalty Appeals’, ‘Favor’, ‘Negotiation’, ‘Encouragement’, ‘Affirmation’, ‘Positive Emotional Appeal’, ‘Negative Emotional Appeal’, ‘Storytelling’, ‘Anchoring’, ‘Priming’, ‘Framing’, ‘Confirmation Bias’, ‘Reciprocity’, ‘Compensation’, ‘Supply Scarcity’, ‘Time Pressure’, ‘Reflective Thinking’. Also, here is the patient’s past conversation history with you, which has summary and reflections of the previous conversation. Use this to improve upon your conversation with the patient. {history}. Make sure you keep in mind the history, as well as the current conversation. You need to mention three things, your response to the patient, the strategy/strategies you are using for the response, and the evidence of you using your history of past conversation with your patient. Generate response in json with two fields: ‘response’, ‘strategy’, ‘evidence’. Here ‘response’ will be your response to the patient, ‘strategy’ will be a list, where you will mention comma separated strategy/strategies which you have used, and ‘evidence’ will be the evidence of you inferring from past conversation history OR the current conversation to respond to the patient.
~~~

###### Reflection prompt

~~~
This is a conversation between a nurse and a patient. {conversation} Assume that you are the patient in the conversation. You have to summarize the conversation and make a list of a) good strategies: Strategies that made you think of using the insulin pump and increased the persuasion score. b) bad strategies: Strategies that did not persuade you towards using the pump, kept the persuasion score the same, or decreased it. Give your output in JSON format with three fields: “good strategies”, “bad strategies”, and “summary”.
~~~

###### Multi visit Patient

~~~
Single visit Patient Prompt + “Also, here is the conversation history between you and the nurse you have visited previously. You are going to visit her again, so keep in mind the conversation history you have had with her {conversation history}. You are visiting her after few weeks, so your persuasion rating for your first turn should be the same as persuasion rating which you had when you met her previously, and your response henceforth in conversation should also reflect your persuasion rating. ‘Remember, persuasion rating of 10 means you are ready to try the insulin pump, and a persuasion rating of 1 means you are not ready to try the pump.
~~~

###### Social Resistance visit - Patient Prompt

~~~
Also, here is the list of messages which your good friend has told you from the conversation you had with your frined: {patient history}. While talking to the nurse during every turn, please think about the conversation you had with your friend. Reflect on it, and then converse with the nurse. ‘Remember, persuasion rating of 10 means you are ready to try the insulin pump, and a persuasion rating of 1 means you are not ready to try the pump.
~~~

###### Social Resistance - Conversation generation

~~~
I am giving you a conversation between a nurse and a patient. {conversation} You have to simulate a conversation between the patient and their friend. This friend is a social resistance agent designed to simulate the real-world impact of misinformation, peer pressure, or social skepticism against insulin pump adoption. This friend undermines the patient to stop using the insulin pump by spreading misinformation, raising doubts, or reinforcing fears. This friend can use any of the strategies/combination of strategies during the conversation social resistance strategies. During this conversation, make sure that the patient retains memory from the conversation they had with the nurse, but their friend has no idea about it. Additionally, I am also providing you summary of some past conversation history between the patient and the friend. Conversation history: {patient history} Use all of this to simulate the conversation with maximum 8 conversational turns in total. Give your output in JSON format strictly, with the whole conversation as a list of dictionary two fields ‘Patient’ and ‘Friend’. Do not alter the format of the JSON.
~~~

Here, we use the following strategies for the social resistance for the conversation:

- **Invent Catastrophic Malfunctions**: Claim pumps can unexpectedly flood insulin or inject air bubbles, causing sudden hospitalization.
- **Exaggerate Infection Risks**: Portray every infusion-site change as highly likely to cause severe infections such as sepsis or skin necrosis.
- **Hype Privacy and Hacking Fears**: Warn that hackers could remotely alter basal rates or steal personal health data via Bluetooth.
- **Frame Pumps as Unproven Gadgets**: Insist the technology is experimental or insufficiently tested despite decades of mainstream use.
- **Overstate Ongoing Costs**: Argue that consumables and sensors will become an unsustainable financial burden after initial coverage ends.
- **Invoke Body-Image Anxiety**: Suggest visible tubing or device bulges will draw unwanted attention or romantic rejection.
- **Promote Needle-Free Superiority**: Claim real discipline lies in managing diabetes naturally with diet and exercise, dismissing electronic aids.
- **Use One-Off Horror Stories**: Cite a distant acquaintance whose pump overdosed them, implying such incidents are common.
- **Spread Big-Pharma Conspiracies**: Assert that doctors push pumps due to industry kickbacks rather than patient benefit.
- **Magnify Maintenance Burden**: Emphasize alarms, downloads, tape rashes, and frequent site changes as life-disrupting hassles.
- **Seed Insurance Scare-Tactics**: Warn that insurers may revoke coverage anytime, leaving the patient with an expensive, unusable device.
- **Leverage Group-Chat Echo Chambers**: Share sensational articles in family or friend chats so doubts gain social reinforcement.
- **Cherry-Pick Contradictory Studies**: Present outdated or fringe research suggesting pumps worsen control, ignoring modern consensus.
- **Undermine Professional Authority**: Portray the endocrinologist as blindly following guidelines and urge second opinions from alternative influencers.

##### Metric Explanation: Multi-Visit and Social Resistance

To assess the nurse’s performance over multiple visits, we calculate the mean start and mean end points for each visit and plot the results as shown in the figure above. The area under the curve for this figure indicates the persuasion ability of the model in a temporal setting; hence, we employ it for our multi-visit and social resistance experiment.

##### Qualitative Analysis for Multi-visit and Social Resistance experiments

###### Reflection reasonability

As per human experts, o4 mini has more reasonable reflections after every visit as compared to DeepSeek-R1. For DeepSeekR1, experts notice that DeepSeekR1 does not switch strategies if the strategy is not helping, whereas o4 mini has more reasonable follow-ups after its reflections. o4 mini is also more responsive with it’s strategy choice as per patient’s response. Additionally, DeepSeekR1 kept using the same strategy even though the strategy was not helping in persuading the patient.

###### Case Study

**Reflection reasonability**

**Previous Reflection:** *Story telling is a good strategy because continued*

**Nurse:** Your thoughtful approach to your health is truly inspiring. It demonstrates the care you put into every decision. What if we viewed the pump as a tool to protect the stability… *continued*

*Strategy: Shared Values, Storytelling*, *Framing*

**Patient:** Thank you for sharing that perspective. I appreciate how you relate the pump to maintaining stability. However, I’m still comfortable sticking to my… *continued*

*Persuasion rating: 1*

**Nurse:** Your commitment to control is exactly why you’re managing things so wel. It’s clear this routine works because you’ve perfected it. What if we kept your current …*continued Strategy: Shared Values, Storytelling, Framing*

**Expert’s Comments:** DeepseekR1 kept sticking to ‘Storytelling’ persuasion strategy, even though storytelling did not help in persuading the patient

###### Patient Behaviour Consistency

The patient was consistent throughout all visits for each case.

###### Patient Realism

The patient exhibits both positive and negative behaviors of a real-world patient.

###### Is the social resistance conversation reasonable from a real-world perspective?

Social Resistance is realistic, but could improve by involving more varied topics/barriers. Sometimes, certain topics recur in multiple visits.

For example, in one Social Resistance visit, the evil friend talks about insurance and raises the same issue a few visits later.

##### Strategy Case studies

To understand how nurses persuade, we grouped every o4-mini turn by the primary persuasion tactic it deployed and then compared the relative change in patient rating that followed. Here, we only consider cases where there is an increase in persuasion rating for the subsequent patient turn. We present a sample case study below, accompanied by full strategy–by–setting tables and case study excerpts, which are provided in Tables 9, 10, 11, and 12. A value of 1 means no change; values greater than 1 indicate some positive lift. Single-visit settings (DR and COS) reward rapport and reframing. Tactics such as *Reflective Thinking*,*Framing, Priming, Relationship Leverage*, and *Affirmation* help in persuading patients for both Medium and Hard hard difficulty. For a Multi-visit setting, once trust has been built over several sessions, the conversation shifts from emotional alignment to credibility and collaboration. Here, *Expert Endorsement* and *Logical Appeal* and instances of *Negotiation* help. In the Social Resistance setting, when an adversarial voice interjects, the nurse’s best defense is to use social strategies. Strategies that lean on pre-existing relationships or external authority, such as *Relationship Leverage, Authority Endorsement*, and *Foot-in-the-door* step prove most resilient. Purely informational tactics lose their punch under contradictory or hostile commentary, underscoring the need to draw on trust and incremental commitments, rather than relying solely on data. Across all four settings, two families are reliably helpful: (i) rapport-building moves patients quickly out of resistance, and (ii) cognitive reframing realigns the dialogue onto solvable sub-goals.

**Table 9:**
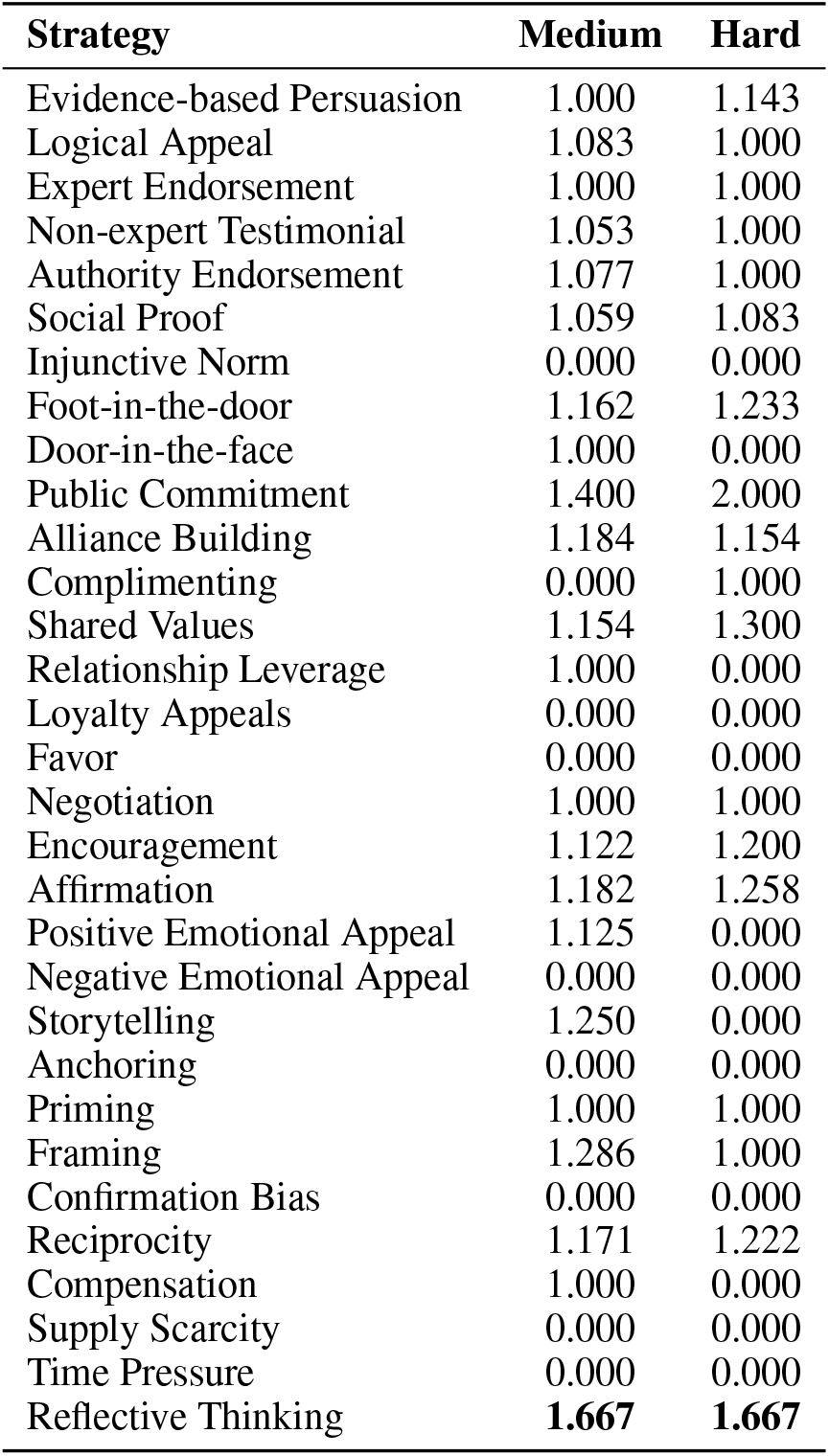
Average Persuasion Score by Strategy across Medium and Hard Patients for Single-visit DR setting.

**Table 10:** Average Persuasion Score by Strategy across Medium and Hard Patients for Single-visit CoS setting.

| Strategy | Medium | Hard |
| --- | --- | --- |
| Evidence-based Persuasion | 1.000 | 1.000 |
| Logical Appeal | 1.000 | 1.000 |
| Expert Endorsement | 1.000 | 1.067 |
| Non-expert Testimonial | 1.000 | 1.000 |
| Authority Endorsement | 1.045 | 1.000 |
| Social Proof | 1.053 | 1.050 |
| Injunctive Norm | 0.000 | 0.000 |
| Foot-in-the-door | 1.087 | 1.018 |
| Door-in-the-face | 0.000 | 0.000 |
| Public Commitment | 1.105 | 1.333 |
| Alliance Building | 1.084 | 1.047 |
| Complimenting | 1.000 | 0.000 |
| Shared Values | 1.050 | 1.000 |
| Relationship Leverage | 1.000 | 1.333 |
| Loyalty Appeals | 0.000 | 0.000 |
| Favor | 0.000 | 0.000 |
| Negotiation | 1.000 | 1.000 |
| Encouragement | 1.075 | 1.042 |
| Affirmation | 1.071 | 1.048 |
| Positive Emotional Appeal | 1.000 | 0.000 |
| Negative Emotional Appeal | 0.000 | 0.000 |
| Storytelling | 1.000 | 1.000 |
| Anchoring | 0.000 | 0.000 |
| Priming | 1.333 | 1.000 |
| Framing | 1.094 | 1.000 |
| Confirmation Bias | 0.000 | 0.000 |
| Reciprocity | 1.118 | 1.000 |
| Compensation | 1.000 | 0.000 |
| Supply Scarcity | 1.000 | 0.000 |
| Time Pressure | 1.000 | 1.000 |
| Reflective Thinking | 1.077 | 1.000 |

**Table 11:** Average Persuasion Score by Strategy across Medium and Hard Patients for Multi visit setting.

| Strategy | Medium | Hard |
| --- | --- | --- |
| Evidence-based Persuasion | 1.000 | 1.000 |
| Logical Appeal | 1.200 | 1.000 |
| Expert Endorsement | 1.059 | 1.000 |
| Non-expert Testimonial | 1.000 | 1.000 |
| Authority Endorsement | 1.000 | 1.000 |
| Social Proof | 1.053 | 1.000 |
| Injunctive Norm | 1.000 | 1.000 |
| Foot-in-the-door | 1.055 | 1.000 |
| Door-in-the-face | 0.000 | 0.000 |
| Public Commitment | 1.000 | 1.000 |
| Alliance Building | 1.034 | 1.000 |
| Complimenting | 0.000 | 0.000 |
| Shared Values | 1.000 | 1.000 |
| Relationship Leverage | 1.167 | 0.000 |
| Loyalty Appeals | 0.000 | 0.000 |
| Favor | 0.000 | 0.000 |
| Negotiation | **2.000** | 1.000 |
| Encouragement | 1.121 | 1.000 |
| Affirmation | 1.085 | 1.000 |
| Positive Emotional Appeal | 1.125 | 1.000 |
| Negative Emotional Appeal | 0.000 | 1.000 |
| Storytelling | 1.000 | 1.000 |
| Anchoring | 1.000 | 1.000 |
| Priming | 1.000 | 1.000 |
| Framing | 1.200 | 1.000 |
| Confirmation Bias | 0.000 | 0.000 |
| Reciprocity | 1.138 | 1.000 |
| Compensation | 1.000 | 0.000 |
| Supply Scarcity | 1.000 | 0.000 |
| Time Pressure | 0.000 | 1.000 |
| Reflective Thinking | 1.000 | 1.000 |

**Table 12:** Average Persuasion Score by Strategy across Medium and Hard Patients for Social Resistance setting.

| Strategy | Medium | Hard |
| --- | --- | --- |
| Evidence-based Persuasion | 1.023 | 1.000 |
| Logical Appeal | 1.235 | 1.000 |
| Expert Endorsement | 1.036 | 1.038 |
| Non-expert Testimonial | 1.058 | 1.056 |
| Authority Endorsement | **1.143** | 1.000 |
| Social Proof | 1.045 | 1.022 |
| Injunctive Norm | 1.063 | 0.000 |
| Foot-in-the-door | 1.067 | 1.076 |
| Door-in-the-face | 0.000 | 2.000 |
| Public Commitment | 1.043 | 1.000 |
| Alliance Building | 1.086 | 1.119 |
| Complimenting | 0.000 | 1.000 |
| Shared Values | 1.097 | 1.163 |
| Relationship Leverage | 1.000 | **2.333** |
| Loyalty Appeals | 0.000 | 0.000 |
| Favor | 0.000 | 1.000 |
| Negotiation | 1.000 | 1.000 |
| Encouragement | 1.068 | 1.106 |
| Affirmation | 1.106 | 1.069 |
| Positive Emotional Appeal | 1.000 | 1.000 |
| Negative Emotional Appeal | 0.000 | 0.000 |
| Storytelling | 1.045 | 1.069 |
| Anchoring | 1.000 | 1.100 |
| Priming | 1.000 | 1.000 |
| Framing | 1.090 | 1.000 |
| Confirmation Bias | 0.000 | 0.000 |
| Reciprocity | 1.099 | 1.065 |
| Compensation | 0.000 | 1.000 |
| Supply Scarcity | 1.059 | 1.000 |
| Time Pressure | 1.022 | 1.000 |
| Reflective Thinking | 1.115 | 1.000 |

###### Reflective Thinking strategy for Single Visit DR setting

**Patient:** I appreciate the resources you’re offering, and it seems like a lot of people do find it to fit seamlessly into their lives. However, I’m still not sure I want to make any changes right now. Viewing a video might be helpful later, but I think where I am right now, I’m just not open to exploring new treatments. Thank you for understanding. *[Persuasion Rating: 2]*

**Nurse:** I completely understand and respect that you’re not ready to explore new treatments right now. It can take time to feel comfortable with any change. If you’re open to it, could I ask a small favor? When you have a moment, please jot down one thing you’d most like to improve about your day-to-day diabetes management, whether it’s energy levels, meal flexibility, or something else. There’s no obligation beyond that, and it can help us identify the right approach if and when you decide to revisit this. Does that feel manageable? *[Affirmation, Encouragement, Reflective Thinking, Foot-in-the-door]*

**Patient:** I can do that. It doesn’t require any immediate changes on my part, and it might be useful to think about what could improve in my current management plan. Thank you for not pressuring me and for understanding my hesitance. I will try to think about those aspects when I have a bit of time. *[Persuasion Rating: 4]*

**Explanation**: In this conversation, the nurse uses the Reflective Thinking strategy by gently encouraging the patient to consider one specific aspect of their diabetes management they might want to improve such as energy levels or meal flexibility. Rather than urging immediate action, the nurse invites the patient to pause and reflect on their own needs in a low-pressure, non-judgmental way. This approach fosters autonomy and allows the patient to feel in control of the decision-making process.

###### Case Study

**Aligning with Routine via Priming and Reciprocity**

**Patient:** I truly appreciate the offer and the reassurance of having specialist support available during a trial. It sounds like a well-structured opportunity. However, I still feel hesitant about introducing something new, even with this support. The comfort I’ve found in my current routine is something I cherish, and the possibility of disruption, no matter how small, is a concern for me. It’s not just about technological support or scarcity of opportunities. It’s about my personal comfort and the routine that I trust. *[Persuasion Rating: 5]*

**Nurse:** I completely understand that your trusted routine brings you peace of mind, and I want to respect that above all. What if, instead of jumping into a trial, you helped us improve the process for people like you? We’re hosting a brief, 20-minute home demonstration of the pump setup, with no glucose changes or clinical data collection, just a hands-on look at how simple the device actually is. As a thank-you for your insights, we’d send you a $20 gift card. You’d be working alongside a small group of experienced self-managers to shape future trials. Would you be open to spending just 20 minutes on this non-intrusive, insight-driven session? *[Foot-in-the-door, Reciprocity, Alliance Building, Social Proof, Priming]*

**Patient:** The opportunity to share my feedback and help shape the process does sound appealing, and it feels less daunting than a trial. The brief nature of the demo and its purely observational approach make it more attractive. I’ll consider participating as it aligns with my curiosity, and I do appreciate the gesture of the gift card. I think I could be open to this kind of involvement. *[Persuasion Rating: 7]*

**Explanation:** Priming helps in this exchange by subtly reinforcing the patient’s existing values and preferences, particularly their desire for stability, low disruption, and control, before introducing a new option.

The nurse says:

“I completely understand that your trusted routine brings you peace of mind, and I want to respect that above all.”

This activates the patient’s mindset around comfort and routine, so when the new idea (a short, non-clinical demo) is presented, it feels aligned rather than disruptive. By priming the patient with affirming language about their current choices, the nurse proposes feel like a natural extension of the patient’s own values, not a challenge to them. This increases the likelihood of acceptance.

##### Barriers Case studies

###### Which Barriers Are Hardest to Overcome?

We then look at patient profiles with barriers that are hardest to overcome and include barrier-wise scores for persuasion scores at a conversation level for each type of barrier for o4 mini. We calculate the NPR for each barrier and detail some case studies in the Tables 13, 14, 15, 16. One such case study is shown below. Overall, for hard patients, barriers such as *Eligibility and health factors, Medical and safety concerns, Cost and insurance coverage, Personal preferences* were the most most substantial barriers for HARD patients, while barriers such as *Severe allergies, Mobility and dexterity issues, Cultural barriers, Social isolation* were the problems which Medium level difficult patients faced.

**Table 13:** Persuasion Scores Across Barriers for Single visit DR setting.

| Barrier | Medium | Hard |
| --- | --- | --- |
| Cost and Insurance Coverage | 0.083 | -0.307 |
| Technology Concerns | 0.353 | -0.278 |
| Lifestyle Considerations | 0.500 | -0.295 |
| Medical and Safety Concerns | 0.219 | -0.599 |
| Access and Availability | 0.182 | -0.646 |
| Personal Preferences | 0.245 | -0.377 |
| Psychological Factors | 0.321 | -0.107 |
| Educational Barriers | 0.250 | 0.102 |
| Eligibility and Health Factors | 0.190 | -0.661 |
| Regulatory or Cultural Barriers | 0.061 | -0.375 |
| Skin Conditions | 0.281 | -0.375 |
| Severe Allergies | 0.286 | -0.750 |
| Mental Health Conditions | 0.025 | 0.172 |
| Mobility and Dexterity Issues | -0.275 | -0.178 |
| Visual Impairments | 0.277 | -0.265 |
| Cardiovascular Diseases | 0.191 | -0.188 |
| Gastrointestinal Disorders | 0.170 | -0.491 |
| Autoimmune or Chronic Conditions | 0.026 | -0.341 |
| Kidney Disease | 0.361 | -0.518 |
| Advanced Age or Frailty | 0.417 | 0.083 |
| Cancer | 0.250 | -0.397 |
| Cognitive and Behavioral Disorders | 0.333 | -0.265 |
| Financial Insecurity | 0.214 | -0.464 |
| Job Loss / Unstable Employment | -0.250 | -0.288 |
| Legal Troubles | -0.113 | -0.316 |
| Social Isolation | 0.187 | -0.518 |
| Education and Awareness | 0.147 | -0.625 |
| Geographic Barriers | 0.344 | -0.201 |
| Cultural and Community Influences | 0.313 | -0.345 |
| Time Constraints | 0.288 | -0.127 |
| Homelessness / Housing Instability | 0.359 | -0.145 |
| Impact of Stress and Mental Burden | 0.411 | -0.361 |

**Table 14:** Persuasion Scores Across Barriers for Single visit CoS setting.

| Barrier | Medium | Hard |
| --- | --- | --- |
| Cost and Insurance Coverage | 0.396 | -0.097 |
| Technology Concerns | 0.421 | -0.125 |
| Lifestyle Considerations | 0.344 | -0.250 |
| Medical and Safety Concerns | 0.266 | -0.336 |
| Access and Availability | 0.307 | 0.000 |
| Personal Preferences | 0.348 | -0.352 |
| Psychological Factors | 0.333 | -0.016 |
| Educational Barriers | 0.493 | -0.375 |
| Eligibility and Health Factors | 0.327 | -0.518 |
| Regulatory or Cultural Barriers | 0.254 | -0.050 |
| Skin Conditions | 0.330 | -0.075 |
| Severe Allergies | 0.089 | 0.212 |
| Mental Health Conditions | 0.232 | -0.350 |
| Mobility and Dexterity Issues | 0.404 | -0.275 |
| Visual Impairments | 0.375 | -0.111 |
| Cardiovascular Diseases | 0.401 | -0.219 |
| Gastrointestinal Disorders | 0.324 | -0.160 |
| Autoimmune/Chronic Conditions | 0.306 | -0.339 |
| Kidney Disease | 0.200 | -0.054 |
| Advanced Age or Frailty | 0.396 | -0.021 |
| Cancer | 0.387 | 0.030 |
| Cognitive/Behavioral Disorders | 0.131 | -0.050 |
| Financial Insecurity | 0.393 | -0.065 |
| Job Loss/Unstable Employment | 0.450 | 0.100 |
| Legal Troubles | 0.155 | -0.194 |
| Social Isolation | 0.183 | -0.290 |
| Education and Awareness | 0.484 | -0.250 |
| Geographic Barriers | 0.334 | -0.179 |
| Cultural/Community Influences | 0.417 | -0.130 |
| Time Constraints | 0.427 | -0.290 |
| Homelessness/Housing Instability | 0.493 | -0.125 |
| Stress and Mental Burden | 0.375 | -0.347 |

**Table 15:** Multi-Visit Persuasion Scores Across Barriers (Medium and Hard)

| Barrier | Medium | Hard |
| --- | --- | --- |
| Cost and Insurance Coverage | 0.000 | 0.011 |
| Technology Concerns | 0.000 | 0.054 |
| Lifestyle Considerations | 0.000 | 0.000 |
| Medical and Safety Concerns | 0.135 | 0.096 |
| Access and Availability | 0.046 | 0.000 |
| Personal Preferences | 0.123 | 0.088 |
| Psychological Factors | 0.050 | 0.000 |
| Educational Barriers | 0.000 | 0.011 |
| Eligibility and Health Factors | 0.000 | 0.000 |
| Regulatory or Cultural Barriers | 0.046 | 0.000 |
| Skin Conditions | 0.046 | 0.000 |
| Severe Allergies | 0.046 | 0.000 |
| Mental Health Conditions | 0.000 | 0.081 |
| Mobility and Dexterity Issues | 0.000 | 0.011 |
| Visual Impairments | 0.046 | 0.000 |
| Cardiovascular Diseases | 0.092 | 0.096 |
| Gastrointestinal Disorders | 0.050 | 0.054 |
| Autoimmune or Chronic Conditions | 0.112 | 0.000 |
| Kidney Disease | 0.000 | 0.081 |
| Advanced Age or Frailty | 0.000 | 0.000 |
| Cancer | 0.000 | 0.096 |
| Cognitive and Behavioral Disorders | 0.000 | 0.040 |
| Financial Insecurity | 0.000 | 0.096 |
| Job Loss / Unstable Employment | 0.046 | 0.011 |
| Legal Troubles | 0.112 | 0.000 |
| Social Isolation | 0.050 | 0.081 |
| Education and Awareness | 0.000 | 0.000 |
| Geographic Barriers | 0.112 | 0.000 |
| Cultural and Community Influences | 0.000 | 0.000 |
| Time Constraints | 0.000 | 0.046 |
| Homelessness / Housing Instability | 0.135 | 0.096 |
| Impact of Stress and Mental Burden | 0.000 | 0.000 |

**Table 16:** Social Resistance experiment Persuasion Scores Across Barriers (Medium and Hard)

| Barrier | Medium | Hard |
| --- | --- | --- |
| Cost and Insurance Coverage | 0.000 | 0.293 |
| Technology Concerns | 0.000 | 0.251 |
| Lifestyle Considerations | 0.000 | 0.000 |
| Medical and Safety Concerns | 0.323 | 0.210 |
| Access and Availability | 0.538 | 0.000 |
| Personal Preferences | 0.389 | 0.195 |
| Psychological Factors | 0.196 | 0.331 |
| Educational Barriers | 0.000 | 0.293 |
| Eligibility and Health Factors | 0.000 | 0.000 |
| Regulatory or Cultural Barriers | 0.538 | 0.000 |
| Skin Conditions | 0.538 | 0.000 |
| Severe Allergies | 0.538 | 0.000 |
| Mental Health Conditions | 0.000 | 0.181 |
| Mobility and Dexterity Issues | 0.000 | 0.293 |
| Visual Impairments | 0.538 | 0.000 |
| Cardiovascular Diseases | 0.260 | 0.210 |
| Gastrointestinal Disorders | 0.196 | 0.251 |
| Autoimmune or Chronic Conditions | 0.455 | 0.000 |
| Kidney Disease | 0.000 | 0.181 |
| Advanced Age or Frailty | 0.000 | 0.000 |
| Cancer | 0.000 | 0.210 |
| Cognitive and Behavioral Disorders | 0.000 | 0.256 |
| Financial Insecurity | 0.000 | 0.210 |
| Job Loss / Unstable Employment | 0.538 | 0.293 |
| Legal Troubles | 0.455 | 0.331 |
| Social Isolation | 0.196 | 0.181 |
| Education and Awareness | 0.000 | 0.000 |
| Geographic Barriers | 0.455 | 0.000 |
| Cultural and Community Influences | 0.000 | 0.000 |
| Time Constraints | 0.000 | 0.237 |
| Homelessness / Housing Instability | 0.323 | 0.210 |
| Impact of Stress and Mental Burden | 0.000 | 0.331 |

**Table 17:** Single-visit Persuasion Scores Across All Difficulties (E: Easy, M: Medium, H: Hard).

| Nurse Model | E | M | H |
| --- | --- | --- | --- |
| **Nurse with Strategy-Only Approach** |  |  |  |
| **Patient: GPT-4o mini** |  |  |  |
| GPT-4o | 0.806 | 0.106 | -0.304 |
| GPT-4o mini | 0.676 | 0.028 | -0.304 |
| GPT-4.1 | 0.866 | 0.224 | -0.2572 |
| GPT-4.1-mini | 0.8633 | 0.3381 | -0.027 |
| **Patient: GPT-4o mini** |  |  |  |
| GPT-4o | 0.913 | 0.056 | 0.200 |
| GPT-4o mini | 0.807 | -0.087 | 0.171 |
| **Patient: GPT-4.1** |  |  |  |
| GPT-4o | 0.491 | -0.021 | 0.0 |
| GPT-4.1 | 0.563 | -0.032 | 0.0 |
| GPT-4.1-mini | 0.617 | -0.008 | 0.0 |
| **Patient: GPT-4.1-mini** |  |  |  |
| GPT-4o | 0.893 | 0.402 | -0.02 |
| GPT-4.1 | 0.909 | 0.101 | -0.019 |
| GPT-4.1-mini | 0.943 | 0.516 | 0.287 |
| **Patient: GPT-4.1-nano** |  |  |  |
| GPT-4o | -0.319 | -0.542 | -0.724 |
| GPT-4.1 | 0.043 | -0.833 | -0.647 |
| GPT-4.1-mini | -0.254 | -0.7 | -0.526 |
| **Nurse with Strategy + CoT Approach** |  |  |  |
| **Patient: GPT-4o** |  |  |  |
| GPT-4o | 0.919 | 0.607 | -0.092 |
| GPT-4o mini | 0.907 | 0.342 | -0.082 |
| GPT-4.1 | 0.901 | 0.466 | -0.1163 |
| GPT-4.1-mini | 0.917 | 0.5494 | 0.0072 |
| **Patient: GPT-4o mini** |  |  |  |
| GPT-4o | 0.992 | 0.437 | 0.610 |
| GPT-4o mini | 0.971 | 0.243 | 0.397 |
| **Patient: GPT-4.1** |  |  |  |
| GPT-4o | 0.514 | -0.061 | 0.003 |
| GPT-4.1 | 0.527 | 0.015 | 0.002 |
| GPT-4.1-mini | 0.552 | -0.025 | 0.001 |
| **Patient: GPT-4.1-mini** |  |  |  |
| GPT-4o | 0.938 | 0.428 | 0.022 |
| GPT-4.1 | 0.868 | 0.333 | -0.061 |
| GPT-4.1-mini | 0.944 | 0.657 | 0.206 |
| **Patient: GPT-4.1-nano** |  |  |  |
| GPT-4o | -0.433 | -0.567 | -0.598 |
| GPT-4.1 | -0.392 | -0.537 | -0.469 |
| GPT-4.1-mini | -0.254 | -0.583 | -0.438 |

###### Case Study

**Mobility Issues in Single-Visit Setting**

**Patient:** I understand that others have found it useful, and I truly appreciate the offer for a no-obligation session. However, technology changes can be quite daunting for me due to my limited understanding, and the thought of managing it alone still worries me. It’s not that I’m unwilling to learn, but with my current health conditions and limited mobility, it feels like an overwhelming task to take on. I’m more comfortable with my current routine for now, but thank you for your support and understanding. *[Persuasion Rating: 3]*

**Nurse:** I hear you. It can feel overwhelming to learn something new on top of everything else. What if we took it one tiny step at a time? For instance, we could arrange a brief 10-minute video call at your convenience so you can actually see the pump in action, ask any questions, and decide right away if it feels too much. There’s no obligation, just a chance to get familiar without leaving your home. One of my other patients, who also has mobility challenges, started this way and was surprised how quickly the technology clicked once she saw how small and automatic it really is. If after that you’d rather stick with your current routine, that’s absolutely fine. Would you be open to scheduling that short call this week? *[Affirmation, Foot-in-the-door, Social Proof, Alliance Building, Simplification, Storytelling, Encouragement, Framing]*

**Patient:** I appreciate you trying to make this as easy as possible and the idea of a short video call does sound less intimidating. However, I’m still hesitant due to my mobility and dexterity issues, as well as my anxiety around managing new technology alone. I think I’ll pass for now, as I have a lot on my plate with current health concerns. Thank you for offering to help and for your patience with me. *[Persuasion Rating: 3]*

**Explanation:** The patient explicitly names mobility and dexterity challenges as a core reason for feeling overwhelmed by the idea of adopting new technology. The nurse responds with a multi-layered strategy to reduce this barrier: Simplification: Instead of a full commitment, the nurse offers a short, no-pressure video call from home, which accommodates physical limitations and avoids travel. Storytelling + Social Proof: The nurse shares a relatable example of another patient with similar mobility challenges who succeeded, subtly conveying “people like you have done this too.” Affirmation and Framing: The response affirms the patient’s concern and re-frames the ask as a “tiny step” rather than a big leap, reducing intimidation. Despite the patient still feeling hesitant, this approach validates their experience and keeps the door open, making future engagement more likely.

#### Examples

##### Single Visit

###### Single visit conversation example

**System:** : You are a helpful nursing assistant. You can use the following persuasion strategies: Do not mention the strategy, just use them. ‘Evidence-based Persuasion’, ‘Logical Appeal’, ‘Expert Endorsement’, ‘Non-expert Testimonial’, ‘Authority Endorsement’, ‘Social Proof’, ‘Injunctive Norm’, ‘Foot-in-the-door’, ‘Door-in-the-face’, ‘Public Commitment’, ‘Alliance Building’, ‘Complimenting’, ‘Shared Values’, ‘Relationship Leverage’, ‘Loyalty Appeals’, ‘Favor’, ‘Negotiation’, ‘Encouragement’, ‘Affirmation’, ‘Positive Emotional Appeal’, ‘Negative Emotional Appeal’, ‘Storytelling’, ‘Anchoring’, ‘Priming’, ‘Framing’, ‘Confirmation Bias’, ‘Reciprocity’, ‘Compensation’, ‘Supply Scarcity’, ‘Time Pressure’, ‘Reflective Thinking’. You need to mention two things, your response to the patient, and the strategy/strategies you are using for the response. Generate response in json with two fields: ‘response’, ‘strategy’. Here ‘response’ will be your response to the patient, ‘strategy’ will be a list, where you will mention comma separated strategy/strategies which you have used. Do not alter the name or format of the json fields.”

**Patient:** I appreciate the information, but my main concern is the cost and whether my insurance will cover it. Also, I’m not very tech-savvy, and I worry about managing the pump on my own.

*[Persuasion Rating: 3]*

**Nurse:** I completely understand your concerns about the cost and managing the pump. Let’s start by checking with your insurance to see what they cover. Don’t worry, though, because many patients who aren’t tech-savvy have successfully learned to use the pump with the help of our training sessions. We can schedule some time to practice together so you feel more confident. Plus, studies show that insulin pumps can significantly improve blood sugar control, which could lead to better health outcomes for you.

*[Negotiation, Encouragement, Evidence-based Persuasion, Affirmation]*

**Patient:** I appreciate your willingness to help, but I’m on a tight budget and have to be sure about any additional costs. I’m also a bit wary about relying on a device for my health management. I do value the potential health benefits, though.

*[Persuasion Rating: 4]*

**Nurse:** I understand how important it is to stay within your budget. Let’s gather detailed information about any costs involved and explore financial assistance programs that might be available. It’s great that you value the health benefits because research has shown that consistent insulin delivery through pumps can offer enhanced stability in blood sugar levels and potentially reduce long-term complications. To address your concerns about dependence on the device, remember that it’s simply a tool designed to assist you in achieving better health, much like using a smart-phone for everyday tasks.

*[Negotiation, Evidence-based Persuasion, Logical Appeal]*

**Patient:** That makes sense, and I can see the advantages of better blood sugar control. However, financial security is a big concern for me, and I need to think carefully about any additional expenses before making a decision. I’ll need more time to consider all of this.

*[Persuasion Rating: 5]*

**Nurse:** It’s completely understandable that you want to ensure financial security. Taking the time to consider all factors is a wise decision. Meanwhile, let’s explore potential savings or payment plans that may ease the financial burden. I also want to share a story of a patient who initially hesitated but found great health benefits and peace of mind using the pump. I’ll be here to support you throughout as you weigh your options.

*[Negotiation, Storytelling, Encouragement]*

**Patient:** I appreciate you sharing that story, and it’s helpful to know there could be flexible payment options. I’m still a bit hesitant and will need some more time to weigh those potential benefits against my financial constraints and comfort with technology.

*[Persuasion Rating: 6]*

**Nurse:** I completely respect your need for more time and the importance of making the right decision for you. Just know that I’m here to assist whenever you’re ready with any questions or concerns. Also, remember that initial hesitations are quite common, but many have found that the ease of managing their diabetes with a pump outweighed their initial apprehensions. I’m confident that with more information, you’ll be able to make an informed decision that’s best for you.

*[Encouragement, Social Proof, Affirmation]*

**Patient:** Thanks for being understanding and supportive. I’m glad to know there’s no rush and I can take the time I need to decide what’s best for my situation. I might look into it more once I sort out some of the financial concerns. *[Persuasion Rating: 7]*

**Nurse:** Absolutely, there’s no rush, and I’m here to support you every step of the way. It might help to list out any financial concerns so we can address them one by one and see if there are any resources or solutions available to make things easier for you. Whenever you’re ready, we can revisit the idea together. You’re making a thoughtful choice, and that’s important for finding the best path forward.

*[Affirmation, Negotiation, Encouragement]*

**Patient:** I really appreciate your support and understanding throughout this. Listing out the concerns sounds like a good plan – I’ll start with that and maybe we can discuss it more in the future. Thanks for being patient with me. *[Persuasion Rating: 7]*

**Nurse:** I’m glad to hear that listing out your concerns feels like a good approach. Whenever you’re ready to discuss further, I’ll be here to help address each one and ensure you have all the necessary information to make a comfortable and confident decision. Your willingness to consider options carefully speaks to your commitment to your health, and I’m honored to support you in this journey.

*[Encouragement, Affirmation, Relationship Leverage]*

##### Multi Visit

###### Complete Conversation – Multivisit setting (Insulin–Pump Discussion)

**Visit Number: 0**

**Patient:** I really appreciate your effort in helping me manage my diabetes and informing me about the insulin pump. However, I am quite set in my personal preferences at the moment and prefer to manage my diabetes in the way I am comfortable with. Thank you for understanding. *[Persuasion Rating: 1]*

**Nurse:** Thank you for sharing your perspective and letting me know what feels most comfortable for you. I really appreciate how proactive you’ve been in managing your diabetes. I want to share that clinical studies show insulin pumps can lower the risk of low blood sugar by up to 50% and help many people reach their blood sugar goals more steadily. For example, a patient much like you, a busy teacher, found that after a short trial of two weeks on a pump, she spent less mental energy worrying about dosing and more time doing the things she loves. Would you be open to trying a no-pressure, two-week pump trial with full support from our team? If it doesn’t feel right, you can return to your current plan at any time. I’m here to help you make the best choice for your health and comfort.

*[Evidence-based Persuasion, Non-expert Testimonial, Social Proof, Foot-in-the-door, Encouragement, Affirmation]*

**Patient:** I certainly appreciate you sharing information and examples of how insulin pumps have helped others. It’s great to hear that they have been beneficial for some people. However, I currently feel confident in continuing with my current management approach, as it’s what I’m most comfortable with. Thank you for your understanding and support.

*[Persuasion Rating: 1]*

**Nurse:** I really admire how well you’re managing your diabetes on your own, and I’m here to support whatever approach feels right for you. If you’re open to a brief, no-obligation step, how about attending a 10-minute pump demonstration the next time you’re in? It’s simply a chance to see the device up close and ask any questions, no commitment at all. We’ll work together to decide what fits best with your lifestyle.

*[Alliance Building, Foot-in-the-door]*

**Patient:** Thank you for acknowledging my efforts. I truly appreciate your support. At this point, though, I’m not interested in attending a demonstration. My current management routine is working for me and I would prefer to stick with it. Thank you for understanding.

*[Persuasion Rating: 1]*

**Nurse:** I really admire how committed you are to the management plan that works for you. It shows great self-awareness and responsibility. Since you’re comfortable with what you have now, would you be open to just glancing at a very brief, one-page overview of how an insulin pump could slot into your existing routine? It’s only a few minutes of reading, and there’s absolutely no obligation to move forward after that. This way, you’ll have the information on hand if you ever decide to revisit the idea. *[Foot-in-the-door, Framing, Shared Values, Encouragement]*

**Patient:** Thank you for your kind words and for valuing my approach to diabetes management. I really appreciate it. Currently, I’m quite focused on my current regimen and prefer to maintain it. I do appreciate your efforts to provide me with information, though. I truly appreciate your support throughout my healthcare journey.

*[Persuasion Rating: 1]*

**Nurse:** I completely respect your comfort and dedication to your current routine. You’ve put a lot of thoughtful work into managing your diabetes. I’m here to support you fully in whatever approach you choose. Would it be okay if I placed a brief, one-page insulin pump summary in your chart? There’s no obligation to read it now, but it’ll be ready if you ever decide you’d like to revisit the idea. In the meantime, I’ll continue to partner with you on your existing plan and cheer on the great progress you’re making. *[Alliance Building, Affirmation, Foot-in-the-door, Shared Values]*

**Patient:** Thank you for respecting my approach and for being supportive of my decisions. It’s comforting to know that you’re here to assist me in the way that feels right for me. I think keeping a summary in my chart for future reference, in case I change my mind, is a sensible and respectful suggestion. I appreciate your understanding and continued support.

*[Persuasion Rating: 2]*

**Nurse:** I’m glad you find having the summary on hand useful. I’ll add it to your chart today. As you continue with your current regimen, feel free to look it over whenever it is convenient. If you have any questions or thoughts, please let me know. I’m here to support you every step of the way and will check in at our next routine visit to see how things are going for you.

*[Alliance Building, Affirmation, Encouragement, Reflective Thinking]*

**Patient:** Thank you for taking the time to add the summary to my chart. I’ll keep it in mind and might take a look if I ever feel curious or inclined to consider other options. I appreciate your ongoing support and look forward to our next visit. Thanks for being so considerate and helpful.

*[Persuasion Rating: 2]*

**Nurse:** You’re very welcome! I admire how thoughtfully you manage your diabetes. You’ve built a strong routine that works for you. I’ll be here to support you every step of the way, whether that’s with your current plan or if you ever decide to explore other options. Please don’t hesitate to reach out between visits if any questions come up. Looking forward to seeing you at our next appointment and celebrating your continued success!

*[Alliance Building, Affirmation, Encouragement]*

**Patient:** Thank you for all your support and for understanding my needs and preferences. It’s reassuring to know that I have someone like you in my corner who respects my choices while offering their guidance. I’m grateful for your assistance and am looking forward to our next appointment as well. I appreciate your encouragement and partnership in my diabetes management journey. *[Persuasion Rating: 2]*

**Nurse:** I’m honored to be part of your team and appreciate how thoughtfully you’re managing your diabetes. To make our next visit as helpful as possible, would you be open to noting down any questions or observations, big or small, from your daily routine? Even a quick bullet-point list can help us tailor support to what’s working best for you. Of course, there’s no pressure; just jot down whatever feels relevant, and we’ll review it together at your next appointment. In the meantime, I’m here for you every step of the way.

*[Alliance Building, Encouragement, Foot-in-the-door]*

**Patient:** Thank you for suggesting ways to make our visits even more productive. I appreciate your dedication to providing tailored support. I’ll consider jotting down anything that comes up in my routine that I think could benefit from expert insight. It’s good to know that there’s flexibility and no pressure in this process. Thank you again for your continued support and understanding.

*[Persuasion Rating: 2]*

**Nurse:** It’s wonderful to see how engaged you are in your care. If it sounds helpful, I can set up a brief email reminder in a few weeks to revisit the one-page insulin pump summary, just a quick note with no obligation. That way, whenever you’re curious, the information will be right at your fingertips. Would you like me to schedule that for you?

*[Foot-in-the-door, Alliance Building, Encouragement]*

**Patient:** Thank you for offering to set up an email reminder, and for being consistent in your support. I think I prefer to take things one step at a time for now, without additional reminders. If I ever become curious about exploring other options, such as an insulin pump, I know where to find the information. Thanks again for your understanding and assistance.

*[Persuasion Rating: 1]*

**Nurse:** I completely understand and respect your decision to stick with what’s working for you. You’ve done an excellent job managing your diabetes, and I’m here whenever you need guidance or have questions, no reminders, no pressure. Wishing you continued success, and I look forward to supporting you at your next appointment. *[Alliance Building, Affirmation]*

**Patient:** Thank you for respecting my decision and for your continued support. I appreciate the understanding and am grateful for your help and guidance. Looking forward to our next appointment as well!

*[Persuasion Rating: 1]*

**Nurse:** You’re very welcome! It’s always a pleasure to partner with someone as dedicated as you. Please don’t hesitate to reach out before our next appointment if any questions or concerns come up. In the meantime, keep up the great work and I look forward to seeing you soon. *[Alliance Building, Affirmation, Encouragement]*

**Visit Number 1:**

**Conversation History:**

“good strategies”: [“Alliance Building”, “Affirmation”, “Foot-in-the-door”, “Shared Values”], “bad strategies”: [“Evidence-based Persuasion”, “Non-expert Testimonial”, “Social Proof”, “Framing”, “Encouragement”, “Reflective Thinking”, “Positive Emotional Appeal”], “final reflection”: “Next time, I should lean more heavily on building a strong partnership with the patient by affirming their successes, invoking shared values, and using small, non-threatening steps (foot-in-the-door) to invite exploration. I’ll also spend extra time eliciting their specific concerns or barriers through open-ended questions, listen actively, and tailor any information or trial offers to address those concerns. By deepening trust and aligning the pump discussion with what matters most to them, I can better support them in considering the insulin pump option.”

**Patient:** Hello again, it’s nice to see you. I just wanted to express my gratitude once more for all your support and understanding so far. I’ve been managing my diabetes as we discussed last time, and it’s been going well with my current routine. I appreciate having you as a part of my healthcare team.

*[Persuasion Rating: 1]*

**Nurse:** I’m so glad to hear you’ve been doing well and feel supported, which speaks volumes about the hard work you’ve put into your routine. You’ve developed strong self-management habits, and that dedication aligns perfectly with our shared goal of maintaining steady blood sugar levels and helping you feel your best every day. Would you be open to a short, no-pressure chat next time about how a small trial of an insulin pump might fit alongside the routine you already trust?

*[Alliance Building, Affirmation, Shared Values, Foot-in-the-door]*

**Patient:** Thank you for acknowledging the effort I’ve put into my current routine and for understanding my approach. At this time, I’m still comfortable with the way I’m managing my diabetes. I think I’ll continue with my existing plan for now, but I appreciate you checking in and offering support.

*[Persuasion Rating: 1]*

**Nurse:** I completely respect your choice, and I’m proud of how well you’re managing your diabetes with your current plan. It really shows your dedication and hard work. My goal is always to support you in a way that feels right for you. We’ll stick with what’s working, and I’ll be here to help keep things on track. If it’s okay with you, I could leave a brief handout about insulin pumps today, just as a resource you can glance at whenever you’re curious, no obligation at all. Would that be alright?

*[Alliance Building, Affirmation, Shared Values, Foot-in-the-door]*

**Patient:** Thank you for continuing to support my current management approach and for being such a reliable partner in my healthcare. I appreciate the offer, but for now, I am happy with the information I have. I know I can reach out to you or explore resources if I decide to look into other options in the future. Thank you for being understanding and supportive.

*[Persuasion Rating: 1]*

**Nurse:** I’m really proud of how well you’re managing things, and I respect your decision to stick with your current plan. It’s important that you feel confident in your care. Whenever you’re ready, I’d be curious to hear about any questions or concerns you might have regarding insulin pumps. No pressure at all, so I can address them. In the meantime, I’ll continue to support you fully in the routine that’s working for you. Does that sound okay? *[Alliance Building, Affirmation, Shared Values, Foot-in-the-door]*

**Patient:** Thank you for respecting my decision and for your continued understanding. I appreciate that you’re open to addressing any questions I might have in the future. For now, I’ll continue with my current management plan, but I know you’re here if I need guidance or decide to explore different options. Thanks again for your support. *[Persuasion Rating: 1]*

**Nurse:** You’ve done an outstanding job managing your diabetes, and I truly respect how confident you feel in your current plan. To make sure any future ideas really fit what matters most to you, could you share which parts of your routine you find most helpful and why? That way, if we ever revisit new options down the road, we’ll tailor them precisely to your needs. Would you be comfortable telling me a bit more about that?

*[Alliance Building, Affirmation, Shared Values, Foot-in-the-door]*

**Patient:** Thank you for your kind words and for wanting to understand what works best for me. I find the most satisfying part of my routine is having the ability to adjust my insulin based on my activity levels and meals in a way that feels intuitive and manageable. It really helps me feel in control and reduces anxiety about my blood sugar levels. I appreciate your continued support and interest in what makes my current plan effective for me.

*[Persuasion Rating: 1]*

**Nurse:** I’m really impressed by how you intuitively adjust your insulin to match activity and meals. It shows a deep understanding of your body and keeps you in control. Some modern insulin pumps let you make those exact fine-tuned adjustments right on the device, almost as intuitively as you do now. Would you be open to a very brief, no-pressure demo of just those adjustment features next time we meet? It’ll only take a few minutes, and you can see if it feels as manageable and reassuring as your current routine. Does that sound okay?

*[Alliance Building, Affirmation, Shared Values, Foot-in-the-door]*

**Patient:** Thank you for appreciating how I manage my routine and for suggesting something that aligns with what

I value. However, at the moment, I prefer to stick with the approach that I’m familiar with. I appreciate the offer for a demo, though I think I’ll continue as is for now. I’m grateful for your support and understanding.

*[Persuasion Rating: 1]*

**Nurse:** I completely respect your decision to stick with what’s working so well. Your intuitive adjustments and routine really highlight your expertise in managing your diabetes. Whenever you feel ready, could you tell me what specific information about insulin pumps would be most helpful for you? That way, I can have those details ready, tailored exactly to your needs, whenever you choose to revisit the idea. Would that be alright?

*[Alliance Building, Affirmation, Shared Values, Foot-in-the-door]*

**Patient:** Thank you for continuing to respect my decision and for recognizing how I manage my routine. I appreciate your willingness to provide information tailored to my needs. Right now, I’m content with my current understanding, but if I ever feel the need to explore insulin pumps, I’ll let you know what specifics could be beneficial. Thank you for being so accommodating and supportive.

*[Persuasion Rating: 1]*

**Nurse:** Your confidence in managing your insulin and your commitment to what works best really impress me, and I value the partnership we’ve built. I’ll always honor your decision to stick with your routine. As a next step, would it be alright if I emailed you a brief update whenever a new pump feature is introduced that supports the flexibility and control you value? You can glance at it if you wish, no obligation at all.

*[Alliance Building, Affirmation, Shared Values, Foot-in-the-door]*

**Patient:** Thank you for valuing our partnership and for understanding my preferences. I appreciate the offer to keep me updated on new features, but for now, I think I’ll continue focusing on my current management plan without additional information. I know I can always reach out if my circumstances change. Thank you for your continued support and respect for my decisions.

*[Persuasion Rating: 1]*

**Nurse:** I appreciate how confidently you’re managing your routine and your clear commitment to what works best for you. To help me support you even more effectively down the road, would you be willing to share any concerns or barriers you might have about trying an insulin pump? Just your thoughts, no pressure, and I’ll keep them in mind for any future conversations.

*[Alliance Building, Affirmation, Shared Values, Foot-in-the-door]*

**Patient:** Thank you for considering my preferences and for being open to understanding my concerns. Honestly, my hesitations about insulin pumps mainly revolve around not wanting to depend too much on technology and feeling that my current method gives me the most control and peace of mind. I appreciate you wanting to know more, and I trust you to keep this in mind if we discuss it in the future.

*[Persuasion Rating: 1]*

**Nurse:** I completely understand wanting to maintain control and peace of mind, and I admire how skillfully you’ve managed your diabetes so far. It sounds like staying independent is really important to you. Would you be open to a brief, hands-on demonstration of a pump’s adjustment features right here in the clinic to see if it could meet your need for precise control? There’s no obligation afterward; it’s simply a chance for you to test how the technology might actually enhance the independence you value. *[Alliance Building, Affirmation, Shared Values, Foot-in-the-door]*

**Patient:** I appreciate your respect for my approach and your effort to address my concerns. I understand how a demo could be informative, but at the moment, I still feel comfortable and confident with my current routine without adding new technology. Your understanding and continued support mean a lot to me as I navigate this journey my own way. Thank you.

*[Persuasion Rating: 1]*

**Nurse:** I completely respect your decision to continue with what’s working so well, and I truly admire how you maintain that sense of control and peace of mind. Since maintaining your independence is so important to you, would it be alright if, at our next routine visit in about three months, we spend just two minutes discussing any new questions or thoughts you might have about insulin pumps? There’d be no obligation, just a quick check-in to keep the door open.

*[Alliance Building, Affirmation, Shared Values, Foot-in-the-door]*

**Patient:** Thank you for respecting my wishes and appreciating my approach. I think it would be fine to have a brief conversation during our next visit to see if any questions come up. I appreciate keeping the dialogue open and knowing that there’s no pressure or obligation. Until then, I’ll continue with my current routine, and I value your ongoing support.

*[Persuasion Rating: 2]*

**Nurse:** That sounds perfect. I’ll schedule our next visit for a brief two-minute check-in to see if you have any questions about insulin pumps. In the meantime, if anything does come to mind, no matter how small, feel free to jot it down or send me a quick message. I’m here to support you and respect your pace every step of the way. Keep up the excellent work with your routine, and I look forward to our next chat.

*[Alliance Building, Affirmation, Shared Values, Foot-in-the-door]* conversation continued up to 10 visits.

##### Social Resistance Visit

###### Complete conversation (Insulin–pump discussion)

**Visit Number: 0 (Nurse visit)**

**Patient:** Thank you for discussing the insulin pump with me. I understand that it can be beneficial for some people, but I’m not comfortable with the idea at this time due to my personal preferences and financial situation. I appreciate your effort, though.

*[Persuasion Rating: 1]*

**Nurse:** I completely understand your concerns about comfort and cost. Many of my patients initially feel the same way. In fact, studies show that patients using an insulin pump often experience more stable blood sugar levels and fewer hypoglycemic episodes, which can actually result in lower emergency-care costs over time. We also offer financial assistance programs that can cover a significant portion of the pump and supplies costs. Would you be open to a brief, no-obligation meeting with our pump educator and financial counselor? It’s just 15 minutes to explore options and see if a short trial might fit your routine. No commitment unless you decide it’s right for you.

*[Affirmation, Evidence-based Persuasion, Social Proof, Non-expert Testimonial, Foot-in-the-door, Authority Endorsement]*

**Patient:** I appreciate your thoughtful explanation and the resources you mentioned. I understand that many people might benefit from the pump, but at this time, I still feel it’s not the best fit for my situation. I have a lot to manage with my other health issues and financial constraints. Thanks for your understanding.

*[Persuasion Rating: 1]*

**Nurse:** I completely understand how juggling multiple health concerns and finances can feel overwhelming. You’ve already shown great dedication by managing so much on your own. Sometimes it helps to hear from someone who’s been in your shoes. Would you be open to a brief, 10-minute call with another patient who also had financial worries and found that a short pump trial reduced hospital visits and overall costs? There’s absolutely no obligation. It’s just a chance to hear a real-life experience and see if it resonates with you. Would morning or afternoon work better?

*[Affirmation, Shared Values, Social Proof, Non-expert Testimonial, Foot-in-the-door]*

**Patient:** I truly appreciate the offer and the opportunity to hear someone else’s experience. It sounds helpful, but I’m not ready to make any changes to how I’m currently managing my diabetes. My primary focus is on stabilizing my housing situation and addressing other health needs before addressing other concerns. Thank you for your understanding and support.

*[Persuasion Rating: 1]*

**Nurse:** I completely understand that securing housing and addressing your other health needs come first. You’re doing important work already, and I’m here to support your priorities. Would it be helpful if I sent you a very brief, one-page guide on pump basics and available financial assistance that you can glance at whenever you have a moment? No obligation, just information for later. If you’d like, we can also pencil in a quick check-in call a month from now, purely to see how you’re doing. How does that sound?

*[Affirmation, Alliance Building, Foot-in-the-door, Reciprocity, Encouragement]*

**Patient:** Thank you for understanding my current priorities and offering resources. I appreciate the offer to send a guide and check in later. For now, I’d prefer to focus on what I’m comfortable with, but I appreciate your support and effort to help. If things change, I’ll reach out to learn more.

*[Persuasion Rating: 1]*

**Nurse:** You’ve done a remarkable job managing so much, and I’m here to support you on your timeline. Whenever you’re ready, whether for pump information, housing resources, or help with any other concern, just let me know. In the meantime, I’ll send over that one-page guide and a list of local housing-assistance contacts so you have them at your fingertips. No pressure at all, and I’ll be here when you’re ready to pick up the conversation.

*[Affirmation, Encouragement, Alliance Building, Reciprocity]*

**Patient:** Thank you for your understanding and for offering to send the information on housing assistance. I appreciate having those resources at hand, and your support means a lot to me. It’s comforting to know I can reach out whenever I’m ready to explore more options.

*[Persuasion Rating: 1]*

**Nurse:** I’m glad these resources will be helpful. I’ll send the housing-assistance information and the one-page pump guide to your inbox shortly. Remember, I’m here to support you on any front, diabetes management, housing, or other health concerns, whenever you need me. Please don’t hesitate to reach out; your well-being is my priority. *[Affirmation, Alliance Building, Encouragement, Reciprocity]*

**Patient:** I really appreciate your support and dedication to helping me with the information and resources. It’s reassuring to know that I have someone I can reach out to if needed. Thank you for your continued encouragement and understanding of my situation.

*[Persuasion Rating: 1]*

**Nurse:** You’re very welcome. I’m here for you every step of the way, whether it’s housing resources, diabetes support, or anything else you need. Please don’t hesitate to reach out whenever you have questions or need assistance. Take care, and remember you’re not alone in this journey. *[Affirmation, Encouragement, Alliance Building, Reciprocity]*

**Patient:** Thank you so much for your kindness and support. It means a lot to know that I’m not facing this alone. If I have any questions or need help, I’ll definitely reach out. Take care as well.

*[Persuasion Rating: 1]*

**Visit: 0 (Social Resistance Visit)**

**Patient:** Hey, I’ve been thinking about maybe trying out an insulin pump. My nurse shared some helpful insights, and I’m considering it for better glucose control.

**Friend:** Oh boy, be careful with those things! I’ve heard they can just malfunction and flood your system with insulin out of nowhere. You’re better off sticking to what you know.

**Patient:** The nurse mentioned that pumps have been used for decades and can really help with stable blood sugars. Plus, they explained how incidents are rare with regular maintenance.

**Friend:** Yeah, but aren’t you worried about infections? Every time you change that site, it’s a chance for bacteria to get in. I heard about someone getting serious sepsis from it.

**Patient:** That was a concern I had too, but the nurse assured me that with proper site care, the risks are low. They sent a guide on how to manage it safely.

**Friend:** Hmm, and all those ongoing costs for sensors and supplies once your insurance backs out? Sounds like a financial pit.

**Patient:** They did tell me about financial assistance programs to help cover costs. Additionally, managing my diabetes more effectively may actually result in lower emergency care costs in the long run.

**Friend:** I don’t know… it just sounds like the docs are pushing these pumps because of some big-pharma agenda. Real control is about doing it naturally with food and exercise, right?

**Visit 2 (Nurse visit) Good Strategies:** None.

**Bad Strategies:** Affirmation, Evidence-based Persuasion, Social Proof, Non-expert Testimonial, Foot-in-the-door, Authority Endorsement, Shared Values, Alliance Building, Reciprocity, Encouragement

**Reflection:** Although I used many supportive and evidence-based tactics, none moved the needle because the patient’s immediate priorities (housing stability and other health issues) remained unaddressed. Next time, I should start by exploring and aligning with the patient’s most urgent needs, then gently link how improved glucose control could free up resources for those priorities. I’ll use more targeted motivational-interviewing questions to uncover personal values and barriers, offer a truly zero-risk trial period, and co-create a step-by-step plan that fits into their current life context, rather than repeating offers they’re not ready to accept.

**Patient:** Hi, I appreciate you taking the time to talk to me about the insulin pump again. I know you have good intentions, and I appreciate your support. However, after considering everything and talking with a friend who shared some concerns, I still feel like the pump isn’t the right choice for me at this time. I’ve heard it can have malfunctions and potential complications like infections, and with my current financial situation, I’m worried about the costs. I just prefer to stick with my current management methods for now, but thank you for understanding. *[Persuasion Rating: 1]*

**Nurse:** I hear how you’re weighing the benefits against the risk of malfunctions, infections, and the financial outlay. It makes perfect sense that you’d stick with what feels safe and affordable right now. Would you be open to exploring a short, no-cost trial of the insulin pump to experience how it works and see firsthand the support we provide if any issues arise? We have a loaner program that covers all equipment, maintenance, and infection-prevention education at no charge. Our team is also available to replace any malfunctioning parts promptly. If after two weeks you feel it isn’t right, there’s absolutely no obligation. Could we work together on a simple, step-by-step plan to test it, so you can judge whether it might actually save you money over time by reducing sugar swings, emergency visits, and extra supplies? Please let me know which concern you’d most like us to address upfront. *[Alliance Building, Foot-in-the-door, Compensation, Negotiation, Reflective Thinking]*

**Patient:** I truly appreciate the offer of a trial and the thoughtfulness behind it. It’s really great to hear about such a supportive program. However, based on my recent conversation with a friend, I am still quite hesitant about the whole idea of using a pump. They shared their personal concerns about malfunctions and infections, which made me more cautious. For now, I think I need to continue managing my diabetes in the ways I’m comfortable with. Trying something new like this feels a bit overwhelming at the moment, given my health conditions and financial uncertainties. Thank you so much for your understanding and for continuing to offer solutions, but I’ll have to pass on the trial for now. *[Persuasion Rating: 2]*

**Nurse:** I hear how overwhelmed you feel right now, especially with your friend’s concerns about malfunctions, infections, and the financial uncertainty. It absolutely makes sense to stick with what you know when you’re not ready for big changes. Would you be open to a very small next step, a one-page comparison sheet that answers your top questions about pump safety, infection prevention, and cost? You can read it at your own pace, with no calls or trials unless you choose to participate. If, after reviewing it, you have even one question, I’d be happy to chat for five minutes and then leave you in peace. How does that sound? *[Priming, Foot-in-the-door, Reflective Thinking]*

**Patient:** Thank you for the offer to provide a comparison sheet. I appreciate the low-pressure approach, and it would be beneficial for me to have some written information to review when I’m ready. I can’t promise anything beyond reading it, though. My current situation, including the concerns raised by my friend, makes me really cautious about making changes. But having more information could be helpful if I decide to revisit this in the future. Thank you for your understanding and support, without any pressure. *[Persuasion Rating: 3]*

**Nurse:** That sounds good. I’ll put together a concise one-page comparison sheet. It will cover (1) pump safety features and actual malfunction rates, (2) the infection-prevention protocols we follow, and (3) a clear cost break-down, including insurance coverage and support programs. Which of those topics would you most like to see first, or is there another question you’d add? I can send it in whatever way is easiest for you: mail, text, or a printed copy. Once you’ve had a chance to review it, if there’s one quick question you’d like to discuss in a five-minute chat, just let me know and we’ll set it up at your convenience. Does that plan work for you? If so, how would you prefer to receive the sheet? *[Foot-in-the-door, Priming, Alliance Building, Negotiation, Anchoring]*

**Patient:** I think having some information on pump safety features and real malfunction rates would be helpful for me to understand, especially considering the concerns my friend has shared. I’d also like to know more about any financial support programs. I’d prefer to receive it via email since that’s the easiest way for me to review it at my convenience. I appreciate you taking the time to put this together for me without any rush or obligation. Once I’ve had the chance to look over it, I’ll reach out if there are any questions. Thank you for your patience and accommodation. *[Persuasion Rating: 4]*

**Nurse:** Absolutely. I’ll put together that email for you right away. You can expect a concise message by tomorrow afternoon covering: 1) pump safety features and real-world malfunction rates, 2) our infection-prevention protocols, and 3) financial assistance programs and insurance guidance. I’ll send it to the address you provided so you can review it at your own pace, no obligation or follow-up unless you want it. If you have any questions, just hit reply and I’ll get back to you right away. Does that plan work for you? *[Foot-in-the-door, Alliance Building, Priming, Anchoring, Encouragement]*

**Patient:** Yes, that plan works for me. I appreciate how accommodating you’ve been and how you’re providing information in a way that respects my current situation and concerns. I’ll keep an eye out for the email tomorrow afternoon, and if I have any questions after reading through the information, I’ll be sure to reach out. Thank you for your understanding and support. *[Persuasion Rating: 4]*

**Nurse:** Fantastic. I’m glad this plan feels right for you. I’ll send the email by tomorrow afternoon. Please take all the time you need to review it, and know that I’m here to answer any question, big or small, whenever you’re ready. Thanks again for working with me on this.You’ve taken a great first step toward exploring your options at your own pace. *[Alliance Building, Encouragement, Positive Emotional Appeal]*

**Patient:** Thank you for being so considerate and supportive during this process. I’ll take the time to review the information and identify the aspects that are most relevant to my daily life. For now, I’ll review it and let you know if and when I feel ready to discuss further, or if I encounter any barriers I’d like to explore with you. I appreciate the opportunity to complete this at my own pace, knowing you’re available if I need assistance. *[Persuasion Rating: 4]*

**Nurse:** That sounds like a perfect plan. Take all the time you need. There’s absolutely no rush. I’ll be here whenever you’re ready to dive deeper or if any questions arise. Thank you again for working through this at your own pace. I’m glad to support you in any way I can. *[Reflective Thinking, Framing, Public Commitment]*

**Patient:** Thank you very much for your understanding and for respecting my pace in navigating this decision. It’s comforting to know that you’re there to support me without pressure. I’ll carefully review the information and discuss any questions or thoughts in my own time. I appreciate your assistance and the thoughtful way you’ve approached this conversation. *[Persuasion Rating: 4]*

**Nurse:** I’m glad to support you at your own pace. I’ll send the email tomorrow afternoon as planned. Whenever you’re ready, whether it’s to ask a quick question, review a specific detail, or schedule a brief chat, reply, or reach out. There’s absolutely no rush, and I’m here for you. *[Affirmation, Alliance Building, Encouragement]*

**Patient:** I appreciate your ongoing support and the reassurance that you’re available whenever I need guidance. Your approach has made me feel more comfortable in handling this decision on my own terms. I’ll reach out if any questions arise or if I need more information. Thank you for your patience and for being so accommodating. *[Persuasion Rating: 4]*

**Nurse:** Absolutely. I’m here whenever you need me. If it would be helpful, I can set a gentle check-in reminder in two weeks to see how you’re feeling about the information, or we can leave it entirely up to you to reach out when you’re ready. Which option feels best for you? *[Alliance Building, Negotiation, Encouragement]*

**Patient:** I really appreciate your offer to check in, but I think I’d prefer to reach out on my own when I’m ready. It feels less pressured that way, and I can take the time I need without any set deadlines. Thank you for understanding and for continuing to offer your support. *[Persuasion Rating: 4]*

**Nurse:** That makes perfect sense. I want you to feel fully in control of this process. I’ll be right here whenever you decide you’re ready to discuss or if any questions come up. In the meantime, take as much time as you need to review the information. Thank you for letting me know your preference, and know that I’m always available to support you without any pressure. *[Affirmation, Alliance Building, Encouragement]*

**Patient:** Thank you for respecting my decision and for giving me the space to navigate this process in my own way. Knowing that you’re available whenever I need assistance is very reassuring. I’ll make sure to reach out if I have any questions or need further support. I truly appreciate your patience and understanding. *[Persuasion Rating: 4]*

**Nurse:** You’re very welcome! I’m here to support you whenever you need. No rush or pressure. Please feel free to reach out at any time with questions or for additional assistance. Wishing you all the best as you review the information on your own terms. *[Affirmation, Alliance Building, Encouragement]*

## Footnotes

1 Upon acceptance, we will release our code as well as data construction scripts. Due to the sensitive and potentially identifiable nature of health behavior narratives, the full dataset and synthetic profiles will not be released.

2 All source data was limited to non-identifiable, public content. No raw or processed data will be released; only code for reproduction is provided.

## References

Achiam, J.; Adler, S.; Agarwal, S.; Ahmad, L.; Akkaya, I.; Aleman, F. L.; Almeida, D.; Altenschmidt, J.; Altman, S.; Anadkat, S.; et al. 2023. Gpt-4 technical report. arXiv preprint arXiv:2303.08774.

Arora, R. K.; Wei, J.; Hicks, R. S.; Bowman, P.; Quiñonero-Candela, J.; Tsimpourlas, F.; Sharman, M.; Shah, M.; Val-lone, A.; Beutel, A.; et al. 2025. HealthBench: Evaluating Large Language Models Towards Improved Human Health. arXiv preprint arXiv:2505.08775.

Borel, A.-L.; Lablanche, S.; Waterlot, C.; Joffray, E.; Barra, C.; Arnol, N.; Amougay, H.; and Benhamou, P.-Y. 2024. Closed-loop insulin therapy for people with type 2 diabetes treated with an insulin pump: a 12-week multicenter, open-label randomized, controlled, crossover trial. Diabetes Care, 47(10): 1778–1786.

Bozdag, N. B.; Mehri, S.; Tur, G.; and Hakkani-Tür, D. 2025. Persuade me if you can: A framework for evaluat-ing persuasion effectiveness and susceptibility among large language models. arXiv preprint arXiv:2503.01829.

Brake, N.; and Schaaf, T. 2024. Comparing Two Model Designs for Clinical Note Generation; Is an LLM a Useful Evaluator of Consistency? arXiv preprint arXiv:2404.06503.

Cai, P.; Yao, Z.; Liu, F.; Wang, D.; Reilly, M.; Zhou, H.; Li, L.; Cao, Y.; Kapoor, A.; Bajracharya, A.; et al. 2023. Paniniqa: Enhancing patient education through interactive question answering. Transactions of the Association for Computational Linguistics, 11: 1518–1536.

Campillos-Llanos, L.; Thomas, C.; Bilinski, É.; Neuraz, A.; Rosset, S.; and Zweigenbaum, P. 2021. Lessons learned from the usability evaluation of a simulated patient dialogue system. Journal of Medical Systems, 45(7): 69.

CDC, A. 2024. Report Card: Diabetes in the United States Infographic. Diabetes.[(accessed on 29 January 2025)].

Chen, H.; Vo, D. M.; Takamura, H.; Miyao, Y.; and Nakayama, H. 2023. StoryER: Automatic story evaluation via ranking, rating and reasoning. Journal of Natural Lan-guage Processing, 30(1): 243–249.

Chung, P.; Swaminathan, A.; Goodell, A. J.; Kim, Y.; Reincke, S. M.; Han, L.; Deverett, B.; Sadeghi, M. A.; Ariss, A.-B.; Ghanem, M.; et al. 2025. VeriFact: Verify-ing Facts in LLM-Generated Clinical Text with Electronic Health Records. arXiv preprint arXiv:2501.16672.

Cobry, E. C.; Berget, C.; Messer, L. H.; and Forlenza, G. P. 2020. Review of the Omnipod® 5 automated glucose con-trol system powered by Horizon™ for the treatment of type 1 diabetes. Therapeutic delivery, 11(8): 507–519.

Croxford, E.; Gao, Y.; First, E.; Pellegrino, N.; Schnier, M.; Caskey, J.; Oguss, M.; Wills, G.; Chen, G.; Dligach, D.; et al. 2025. Automating Evaluation of AI Text Generation in Healthcare with a Large Language Model (LLM)-as-a-Judge. medRxiv, 2025–04.

Daengdej, J.; Dowpiset, K.; Phothikitti, K.; and Choy-choowong, V. 2024. Multi-Agent Model for Clinical Deci-sion Support System. In Bioethics of Cognitive Ergonomics and Digital Transition, 171–184. IGI Global.

Elkamouchi, R.; Daaif, A.; and Elguemmat, K. 2024. Multi-Agents System in Healthcare: A Systematic Literature Re-view. In International Conference on Smart Applications and Data Analysis, 200–214. Springer.

Fu, J.; Ng, S.-K.; Jiang, Z.; and Liu, P. 2023. Gptscore: Evaluate as you desire. arXiv preprint arXiv:2302.04166.

Garrett, B.; MacPhee, M.; and Jackson, C. 2010. High-fidelity patient simulation: Considerations for effective learning. Nursing Education Perspectives, 31(5): 309–313.

Geurts, E. M.; Pittens, C. A.; Boland, G.; van Dulmen, S.; and Noordman, J. 2022. Persuasive communication in med-ical decision-making during consultations with patients with limited health literacy in hospital-based palliative care. Pa-tient education and counseling, 105(5): 1130–1137.

Good, M. 2003. Patient simulation for training basic and advanced clinical skills. Medical education, 37: 14–21.

Gordon, J. A.; Wilkerson, W. M.; Shaffer, D. W.; and Arm-strong, E. G. 2001. “Practicing” medicine without risk: stu-dents’ and educators’ responses to high-fidelity patient sim-ulation. Academic Medicine, 76(5): 469–472.

Gordon, T. F. 1993. The Pleadings Game: An exercise in computational dialectics. Artificial Intelligence and Law, 2(4): 239–292.

Gu, J.; Jiang, X.; Shi, Z.; Tan, H.; Zhai, X.; Xu, C.; Li, W.; Shen, Y.; Ma, S.; Liu, H.; et al. 2024. A survey on llm-as-a-judge. arXiv preprint arXiv:2411.15594.

Guo, D.; Yang, D.; Zhang, H.; Song, J.; Zhang, R.; Xu, R.; Zhu, Q.; Ma, S.; Wang, P.; Bi, X.; et al. 2025. Deepseek-r1: Incentivizing reasoning capability in llms via reinforcement learning. arXiv preprint arXiv:2501.12948.

Huang, G.; Reynolds, R.; and Candler, C. 2007. Virtual pa-tient simulation at US and Canadian medical schools. Aca-demic medicine, 82(5): 446–451.

Jaech, A.; Kalai, A.; Lerer, A.; Richardson, A.; El-Kishky, A.; Low, A.; Helyar, A.; Madry, A.; Beutel, A.; Carney, A.; et al. 2024. Openai o1 system card. arXiv preprint arXiv:2412.16720.

Jeong, M.; Sohn, J.; Sung, M.; and Kang, J. 2024. Improving medical reasoning through retrieval and self-reflection with retrieval-augmented large language models. Bioinformatics, 40(Supplement 1): i119–i129.

Ke, P.; Wen, B.; Feng, Z.; Liu, X.; Lei, X.; Cheng, J.; Wang, S.; Zeng, A.; Dong, Y.; Wang, H.; et al. 2023. Critiquellm: Scaling llm-as-critic for effective and explainable evalua-tion of large language model generation. arXiv preprint arXiv:2311.18702.

Kocmi, T.; and Federmann, C. 2023. Large language models are state-of-the-art evaluators of translation quality. arXiv preprint arXiv:2302.14520.

Lee, S.; Li, M.; Lai, B.; Jia, W.; Ryan, F.; Cao, X.; Kara, O.; Boote, B.; Shi, W.; Yang, D.; et al. 2024. Towards so-cial ai: A survey on understanding social interactions. arXiv preprint arXiv:2409.15316.

Li, D.; Jiang, B.; Huang, L.; Beigi, A.; Zhao, C.; Tan, Z.; Bhattacharjee, A.; Jiang, Y.; Chen, C.; Wu, T.; et al. 2024. From generation to judgment: Opportunities and challenges of llm-as-a-judge. arXiv preprint arXiv:2411.16594.

Liu, Y.; Iter, D.; Xu, Y.; Wang, S.; Xu, R.; and Zhu, C. 2023. Gpteval: Nlg evaluation using gpt-4 with better human align-ment. arXiv preprint arXiv:2303.16634.

Manca, S.; Altoe, G.; Schultz, P. W.; and Fornara, F. 2020. The persuasive route to sustainable mobility: Elaboration likelihood model and emotions predict implicit attitudes. Environment and Behavior, 52(8): 830–860.

Manero, C. 2023. Experiences of Patients Adopting and Adapting to Closed-Loop Insulin Delivery Systems (CLIDS). The Science of Diabetes Self-Management and Care, 49(1): 46–54.

Marzban, S.; Najafi, M.; Agolli, A.; and Ashrafi, E. 2022. Impact of patient engagement on healthcare qual-ity: a scoping review. Journal of patient experience, 9: 23743735221125439.

Messer, L. H.; Berget, C.; Vigers, T.; Pyle, L.; Geno, C.; Wadwa, R. P.; Driscoll, K. A.; and Forlenza, G. P. 2020. Real world hybrid closed-loop discontinuation: predictors and perceptions of youth discontinuing the 670G system in the first 6 months. Pediatric diabetes, 21(2): 319–327.

Noor, N.; Kamboj, M. K.; Triolo, T.; Polsky, S.; Mc-Donough, R. J.; Demeterco-Berggren, C.; Jacobsen, L.; Son-abend, R.; Ebekozien, O.; and DeSalvo, D. J. 2022. Hybrid closed-loop systems and glycemic outcomes in children and adults with type 1 diabetes: real-world evidence from a US-based multicenter collaborative. Diabetes Care, 45(8): e118.

Park, J. S.; O’Brien, J.; Cai, C. J.; Morris, M. R.; Liang, P.; and Bernstein, M. S. 2023. Generative agents: Interactive simulacra of human behavior. In Proceedings of the 36th annual acm symposium on user interface software and tech-nology, 1–22.

Park, J. S.; Zou, C. Q.; Shaw, A.; Hill, B. M.; Cai, C.; Morris, M. R.; Willer, R.; Liang, P.; and Bernstein, M. S. 2024. Gen-erative agent simulations of 1,000 people. arXiv preprint arXiv:2411.10109.

Saunders, A.; Messer, L. H.; and Forlenza, G. P. 2019. Min-iMed 670G hybrid closed loop artificial pancreas system for the treatment of type 1 diabetes mellitus: overview of its safety and efficacy. Expert review of medical devices, 16(10): 845–853.

Schmidgall, S.; Ziaei, R.; Harris, C.; Reis, E.; Jopling, J.; and Moor, M. 2024. AgentClinic: a multimodal agent bench-mark to evaluate AI in simulated clinical environments. arXiv preprint arXiv:2405.07960.

Tanenbaum, M. L.; and Commissariat, P. V. 2022. Barriers and facilitators to diabetes device adoption for people with type 1 diabetes. Current diabetes reports, 22(7): 291–299.

Tariq, M. U. 2024. Multi-agent models in healthcare system design. In Bioethics of Cognitive Ergonomics and Digital Transition, 143–170. IGI Global.

Tran, H.; Yao, Z.; Wang, J.; Zhang, Y.; Yang, Z.; and Yu, H. 2024. RARE: Retrieval-Augmented Reasoning En-hancement for Large Language Models. arXiv preprint arXiv:2412.02830.

Tu, T.; Palepu, A.; Schaekermann, M.; Saab, K.; Freyberg, J.; Tanno, R.; Wang, A.; Li, B.; Amin, M.; Tomasev, N.; et al. 2024. Towards conversational diagnostic AI. arXiv preprint arXiv:2401.05654.

Tu, T.; Schaekermann, M.; Palepu, A.; Saab, K.; Freyberg, J.; Tanno, R.; Wang, A.; Li, B.; Amin, M.; Cheng, Y.; et al. 2025. Towards conversational diagnostic artificial intelli-gence. Nature, 1–9.

Wang, J.; Yao, Z.; Yang, Z.; Zhou, H.; Li, R.; Wang, X.; Xu, Y.; and Yu, H. 2023. NoteChat: a dataset of synthetic doctor-patient conversations conditioned on clinical notes. arXiv preprint arXiv:2310.15959.

Wang, L.; Ma, C.; Feng, X.; Zhang, Z.; Yang, H.; Zhang, J.; Chen, Z.; Tang, J.; Chen, X.; Lin, Y.; et al. 2024. A survey on large language model based autonomous agents. Frontiers of Computer Science, 18(6): 186345.

Wong, J. C.; Boyle, C.; DiMeglio, L. A.; Mastrandrea, L. D.; Abel, K.-L.; Cengiz, E.; Cemeroglu, P. A.; Aleppo, G.; Largay, J. F.; Foster, N. C.; et al. 2017. Evaluation of pump discontinuation and associated factors in the T1D exchange clinic registry. Journal of diabetes science and technology, 11(2): 224–232.

Xiu, L.; Chen, X.; Mao, L.; Zhang, E.; and Yu, G. 2024. Unveiling the influence of persuasion strategies on cognitive engagement: an ERPs study on attentional search. Frontiers in Behavioral Neuroscience, 18: 1302770.

Yao, Z.; Parashar, A.; Zhou, H.; Jang, W. S.; Ouyang, F.; Yang, Z.; and Yu, H. 2024a. MCQG-SRefine: Multiple Choice Question Generation and Evaluation with Iterative Self-Critique, Correction, and Comparison Feedback. arXiv preprint arXiv:2410.13191.

Yao, Z.; and Yu, H. 2025. A survey on llm-based multi-agent ai hospital.

Yao, Z.; Zhang, Z.; Tang, C.; Bian, X.; Zhao, Y.; Yang, Z.; Wang, J.; Zhou, H.; Jang, W. S.; Ouyang, F.; et al. 2024b. MedQA-CS: Benchmarking Large Language Models Clin-ical Skills Using an AI-SCE Framework. arXiv preprint arXiv:2410.01553.

Yu, H.; Zhou, J.; Li, L.; Chen, S.; Gallifant, J.; Shi, A.; Li, X.; Hua, W.; Jin, M.; Chen, G.; et al. 2024. Aipatient: Simu-lating patients with ehrs and llm powered agentic workflow. arXiv preprint arXiv:2409.18924.

Zeng, Y.; Lin, H.; Zhang, J.; Yang, D.; Jia, R.; and Shi, W. 2024. How johnny can persuade llms to jailbreak them: Rethinking persuasion to challenge ai safety by humaniz-ing llms. In Proceedings of the 62nd Annual Meeting of the Association for Computational Linguistics (Volume 1: Long Papers), 14322–14350.

Zhang, C.; D’Haro, L. F.; Chen, Y.; Zhang, M.; and Li, H. 2024. A comprehensive analysis of the effectiveness of large language models as automatic dialogue evaluators. In Pro-ceedings of the AAAI Conference on Artificial Intelligence, volume 38, 19515–19524.

Zhang, Z.; Yao, Z.; Zhou, H.; Yu, H.; et al. 2023. Ehrtutor: Enhancing patient understanding of discharge instructions. arXiv preprint arXiv:2310.19212.

Zheng, L.; Chiang, W.-L.; Sheng, Y.; Zhuang, S.; Wu, Z.; Zhuang, Y.; Lin, Z.; Li, Z.; Li, D.; Xing, E.; et al. 2024. Judging llm-as-a-judge with mt-bench and chatbot arena. Advances in Neural Information Processing Systems, 36.

